# Quantitation of SARS-CoV-2 neutralizing antibodies with a virus-free, authentic test

**DOI:** 10.1101/2021.12.29.21268487

**Authors:** Johannes Roessler, Dagmar Pich, Manuel Albanese, Paul R. Wratil, Verena Krähling, Johannes Christian Hellmuth, Clemens Scherer, Michael von Bergwelt-Baildon, Stephan Becker, Oliver T. Keppler, Alain Brisson, Reinhard Zeidler, Wolfgang Hammerschmidt

## Abstract

Neutralizing antibodies (NAbs), and their concentration in sera of convalescents and vaccinees are a solid correlate of protection from COVID-19. The antibody concentrations in clinical samples that neutralize SARS-CoV-2 are difficult and very cumbersome to assess with conventional virus neutralization tests (cVNTs), which require work with the infectious virus and biosafety level 3 containment precautions. Alternative virus neutralization tests currently in use are mostly surrogate tests based on direct or competitive ELISA formats or use viral vectors with the spike protein as the single structural component of SARS-CoV-2. To overcome these obstacles, we developed a virus-free, safe and very fast (4.5 h) *in vitro* diagnostic test based on engineered yet authentic SARS-CoV-2 virus-like-particles (VLPs). They share all features of the original SARS-CoV-2 but lack the viral RNA genome and thus are non-infectious. NAbs induced by infection or vaccination, but also potentially neutralizing monoclonal antibodies can be reliably quantified and assessed with ease and within hours with our test, because they interfere and block the ACE2-mediated uptake of VLPs by recipient cells. Results from the VLP neutralization test (VLPNT) show excellent correlation to a cVNT with fully infectious SARS-CoV-2 and allow to estimate the reduced neutralization capacity of COVID-19 vaccinee sera with variants of concern of SARS-CoV-2.

**Author summary:** The current pandemic caused by SARS-CoV-2 is a major challenge not only for COVID-19 patients, medical staff, healthcare systems and the general public, but also virologists and clinical laboratories. A particular challenge are safety issues which require biological safety level 3 to work with and study the pathogen. An alternative are virus-like particles (VLPs) of SARS-CoV-2, which are authentic in terms of viral structure and function but are harmless bioproducts in nature. We engineered VLPs which are close-to-perfect mimics of SARS-CoV-2 by all structural, biochemical, physical and functional criteria tested. SARS-CoV-2 VLPs were used in virus neutralization tests (VNTs). Because high concentrations of neutralizing antibodies correlate with protection from COVID-19 practical VNTs are urgently needed. We developed an authentic, virus-free, thus safe yet very fast *in vitro* diagnostic test with SARS-CoV-2 VLPs. Virus neutralizing antibodies induced by natural infection or vaccination but also certain monoclonal antibodies inhibit VLP fusion with recipient cells carrying ACE2. Quantitative results from a conventional neutralization test with fully infectious SARS-CoV-2 and results from the VLP-based neutralization test correlate perfectly. The setup of the test is very flexible and allows to analyze sera for their neutralizing capacity against different variants of concern and in a standardized assay format.

## Introduction

In December 2019 a novel respiratory infectious disease, emerging in the city of Wuhan, China, and leading to an outbreak of severe cases of pneumonia (Wu et al., 2020) marked the beginning of the ongoing pandemic caused by the severe acute respiratory syndrome coronavirus 2 (SARS-CoV-2), a virulent member of the *Coronaviridae* family. The virus likely originated from a wildlife reservoir in bats but spreads easily among humans via droplets and aerosols. As of November 2021, the associated Coronavirus disease 2019 (COVID-19) accounts for more than 5 million deaths worldwide (WHO dashboard, https://covid19.who.int). Aside from subclinical infections, COVID-19 can vary from weak symptoms to mild or severe pneumonia with dyspnea, to critical clinical courses with acute respiratory distress syndrome (ARDS) requiring external ventilation and intensive care.

SARS-CoV-2 is a non-segmented, enveloped *Betacoronavirus* with a positive-sense single-stranded RNA genome of almost 30 kb with open reading frames (ORFs) encoding a replicase polyprotein (ORF1a/ORF1b), four structural proteins spike (S), envelope (E), membrane (M) and nucleoprotein (N, also known as nucleocapsid) and seven accessory proteins (Hu et al., 2021; Mittal et al., 2020). In detail, S is a class I fusion protein, which assembles in homotrimers, comprising three S1 domains on top of three S2 units, each separated by a S1/S2 furin cleavage site. While S1 contains the receptor binding domain (RBD), S2 bears the fusion peptide and two heptad repeats, mediating membrane proximity and fusion (Hofmann and Pohlmann, 2004). Proteolytic processing at the S1/S2 site, which often occurs during egress from a virus producing cell primes S for an additional second cleavage at the S2’ site within the S2 domain. The second cleavage facilitates presentation of the fusion peptide upon endosomal uptake by the susceptible host cell (Shang et al., 2020; Stevens et al., 2021).

Angiotensin-converting enzyme 2 (ACE2) on target cells serves as cellular receptor for SARS-CoV-2, which attaches to ACE2 via the receptor binding motif (RBM) in the S1 subunit. To make the RBM accessible, S in its pre-fusion conformation undergoes conformational changes, exposing one RBD of the protomer in the ACE2-accessible “up” orientation (Mittal et al., 2020). Receptor binding then triggers endocytosis and proteolytic processing of S at the S2’ site by either the cell surface protease TMPRSS2 (Hoffmann et al., 2020a) or endosomal Cathepsin L (CTSL) (Stevens et al., 2021), enabling insertion of the fusion peptide in the host membrane and membrane fusion. Subsequent release of the viral RNA cargo into the cytoplasm initiates translation and further steps downstream to turn the cell into a virus shedding factory (Mittal et al., 2020).

Immunity to SARS-CoV-2 is granted by the innate, but also the adaptive immune system in the form of anti-S antibodies (Hu et al., 2021) causing direct neutralization of virions as well as Fc mediated antibody-dependent cellular phagocytosis (ADCP), complement-dependent cytotoxicity (CDC) and antibody-dependent cellular cytotoxicity (ADCC) (Zohar and Alter, 2020). Besides humoral immunity, cellular immune response also supports broad and durable immune protection, primarily in the form of CD8^+^ T cells targeting the nucleoprotein, but also spike specific CD4^+^ T cells (Cohen et al., 2021; Niessl et al., 2021).

In the course of the pandemic, several mutations in the spike gene led to enhanced viral infectivity and spread. The first major mutation was the single amino acid mutation D614G, which led to higher viral loads and worldwide spread displacing the original Wuhan-2019 strain within only a couple of weeks (Korber et al., 2020). Meanwhile, other strains such as B.1.1.7, the Alpha variant of concern (VOC) also led to enhanced transmissibility (Davies et al., 2021), which is probably due to the N501Y mutation in S, enhancing ACE2 affinity (Zahradnik et al., 2021). B.1.617.2, the Delta-VOC currently predominates as it is more transmissible than the Alpha strain (Burki, 2021; Liu and Rocklov, 2021), yet it is controversial whether this is caused by increased viral loads (Luo et al., 2021; Singanayagam et al., 2021), but the risk to become hospitalized when infected with the Delta-VOC is increased by a factor of about 2 (Sheikh et al., 2021). Moreover, new VOCs such as ‘Omicron’ are a lingering threat.

As a result of the global effort in COVID-19 vaccine development, two mRNA vaccines, BNT162b2 by BioNTech/Pfizer and mRNA-1273 by Moderna, as well as a chimpanzee adenovector ChAdOx1 vaccine (AZD1222) by AstraZeneca were licensed by the FDA and EMA (among other vaccines). For infections with the Alpha- and Delta-VOCs, these vaccines significantly reduce the risk for symptomatic COVID-19, effectively attenuate disease severity and reduce the rate of mortality (Abu-Raddad et al., 2021; Lopez Bernal et al., 2021; Sheikh et al., 2021). The current vaccines, however, do not seem to confer sterile immunity as break-through infections even in fully vaccinated individuals can occur as early as two to three months after the second shot (Levine-Tiefenbrun et al., 2021; Singanayagam et al., 2021).

Neutralizing antibodies (NAbs), induced by infection or vaccination or applied in the form of monoclonal antibodies (mAbs) or convalescent plasma (DeFrancesco, 2020) have the potential to establish immunity to SARS-CoV-2 and to protect from severe COVID-19. As such, the concentration of NAbs is generally accepted as a relevant correlate of protection. In line, high NAb titers were shown to be directly associated with a lower risk of symptomatic SARS-CoV-2 infections and therefore are highly predictive to protect from COVID-19 (Earle et al., 2021; Feng et al., 2021; Khoury et al., 2021). Noteworthy, also anti-S IgG and anti-RBD IgG antibodies, which do not necessarily inhibit viral infection *in vitro*, showed acceptable correlation with protection, yet NAb titers determined with a conventional virus neutralization test (cVNT) were found to correlate best (Feng et al., 2021).

Reliable quantification of NAbs in clinical samples is problematic for several reasons. One reason lies in the diversity of VNTs and their standardization and validation (Krammer, 2021). Undisputed ‘gold standard’ for quantitating NAbs are conventional virus neutralization tests (cVNTs). They rely on replication competent virus stocks, which, as such, guarantee correct virion composition and an authentic infection process. Different versions of cVNTs are in use (Khoury et al., 2020): (i) In multi-cycle assays, NAbs interfere with viral infection and reduce its replication rate, monitored by quantitating the amount of viral antigen generated within a defined period of time post infection. (ii) cVNTs based on limiting dilution use the initial inactivation of the inoculum by NAbs and the reduction of the viral cytopathic effect (CPE) as a function of replica plating of the virus-antibody mix. (iii) Plaque reduction neutralization tests (PRNTs) are based on single infected cells, which give rise to a single plaque, a localized CPE in monolayers of immobilized gel-embedded cells. cVNTs and PRNTs depend on the formation of CPEs and require visual enumeration. As a consequence, the tests are hardly scalable and cumbersome to standardize between different laboratories. All cVNTs involve handling of infectious virus, which, in case of SARS-CoV-2 require typical containment measures of a BSL-3 facility. In addition, all cVNTs take up to several days until readout. Consequently, several alternatives to measure NAb concentration have been developed.

Surrogate virus neutralization tests (sVNT), often performed in an ELISA format, do not specifically quantitate NAbs but antibodies that bind to spike RBD epitopes or antibodies that interfere with the RBD-ACE2 interaction. Given the apparent limitations of sVNTs, pseudotyped virus neutralization tests (pVNTs) have been developed. They quantitate NAbs using susceptible ACE2-positive target cells but rely on viral vectors with spike as the only SARS-CoV-2-derived component. These genetically engineered viral vectors are either replication incompetent, or single-cycle infectious retro-, rhabdo-or lentivirus particles, which contain the spike protein in their envelopes. Infection of suitable cells with these vectors is monitored by *de novo* transcription and translation of a phenotypic reporter protein such as luciferase or a fluorescent protein (Khoury et al., 2020). pVNTs, which require BLS2 laboratory standards, are difficult to reproduce, poorly suitable for up-scaling and automation and their readout takes two to three days. Only recently, a version of pVNT has been proposed, which makes use of a fast lentivirus-based transfer of an enzyme reporter (Miyakawa et al., 2020). Commonly, pVNTs are in wide use but the assembly, morphogenesis, structure and composition of retro-or lentiviral vector particles differ dramatically from that of coronaviruses. The versions of different cVNTs and pVNTs to analyze SARS-CoV-2 NAbs have recently been summarized by Khoury *et al*. (Khoury et al., 2020).

Given the clinical relevance of NAbs and the problems with and limitations of the various VNTs, we developed a virus- and GMO-free diagnostic VNT that is safe and quick and quantifies SARS-CoV-2 NAbs at a level and quality comparable to a cVNT. We established a protocol to produce authentic virus-like-particles (VLPs), which also encompass an activator peptide to trace them. The identity of the VLPs was thoroughly examined by cryo-electron microscopy (cryo-EM) as well as with biochemical methods to validate their biochemical, physical and functional characteristics in comparison to infectious SARS-CoV-2. Our results document that SARS-CoV-2 VLPs enter target cells via ACE2, mediate membrane fusion in endosomes and deliver their luminal protein cargo into the cytosol, thus mimicking all steps of infection of the pathogen prior to viral transcription. Therefore, NAbs that provide protective immunity from SARS-CoV-2 also prevent ‘infection’ with SARS-CoV-2 VLPs. Our test quantitates them and its results show excellent correlation to a cVNT with infectious virus using a set of double-blinded COVID-19 patient serum samples. Additionally, our work demonstrates that this VLP neutralization test (VLPNT) allows for the evaluation of VOCs by simply adapting the assay to the B.1.617.2 Delta variant and measuring reduced NAb titers in sera from COVID-19 vaccinees.

By meeting important requirements for quality, automation, reproducibility, and rapidness, this test is a valuable tool for vaccine and therapeutic antibody development, as well as for high-throughput screening of viral entry inhibitors. As the test format is flexible it can easily be adapted to mutants of SARS-CoV-2 that may emerge in the future.

## Results

### Manufacturing of SARS-CoV-2 VLPs

SARS-CoV-2 VLPs, termed S^+^ VLPs, were generated by transient co-transfection of expression plasmids encoding all four structural proteins of the virus: S (Wuhan-2019, D614G or B.1.617.2), M, N and E in defined stoichiometry into HEK293T cells. To trace the S^+^ VLPs, a fifth expression plasmid was co-transfected to express a chimeric reporter protein consisting of the human CD63 tetraspanin protein and, at its carboxy terminus, an activator of split-nano-luciferase (CD63∼HiBiT). Three days after transfection, assembled S^+^ VLPs were present in large quantities in the cell culture medium. Further purification and concentration were optional and applied if needed to characterize the S^+^ VLPs in detail.

### cryoEM of SARS-CoV-2 VLPs

Cryo-electron microscopy (cryo-EM) of the S^+^ VLPs revealed spherically shaped vesicles in the range of 60 to 150 nm in diameter with a membrane consisting of an intact lipid bilayer (Fig. 1). Like SARS-CoV-2 virions (Ke et al., 2020; Liu et al., 2020; Yao et al., 2020), the VLPs displayed a characteristic corona of dense, needle-like radial proteins protruding perpendicularly from the membrane. On the distal ends of the protrusions spacious heads sit on slimmer stems, suggesting that these structures correspond to the viral glycoprotein spike of SARS-CoV-2. The shape and dimensions of the protrusions which are about 25 nm in length and have a stem width of 7 nm clearly support this assumption. In addition, elongated structures are observed on certain spike bearing particles (white arrows in Fig. 1). These structures might correspond to spike protein protrusions lying down on the vesicle surface, likely caused by surface tension effects prior to plunging of the sample in cryogen. Also, evaporation might reduce the height of the liquid film causing a partial air contact of the particle’s envelope and a redistribution and flattening of surface components in contrast to spikes from the periphery, which maintain their integrity in the surrounding liquid phase. Alternatively, these elongated structures may also correspond to some elements such as fibrous proteins present in the lumen of S^+^ VLPs.

**Figure 1.**
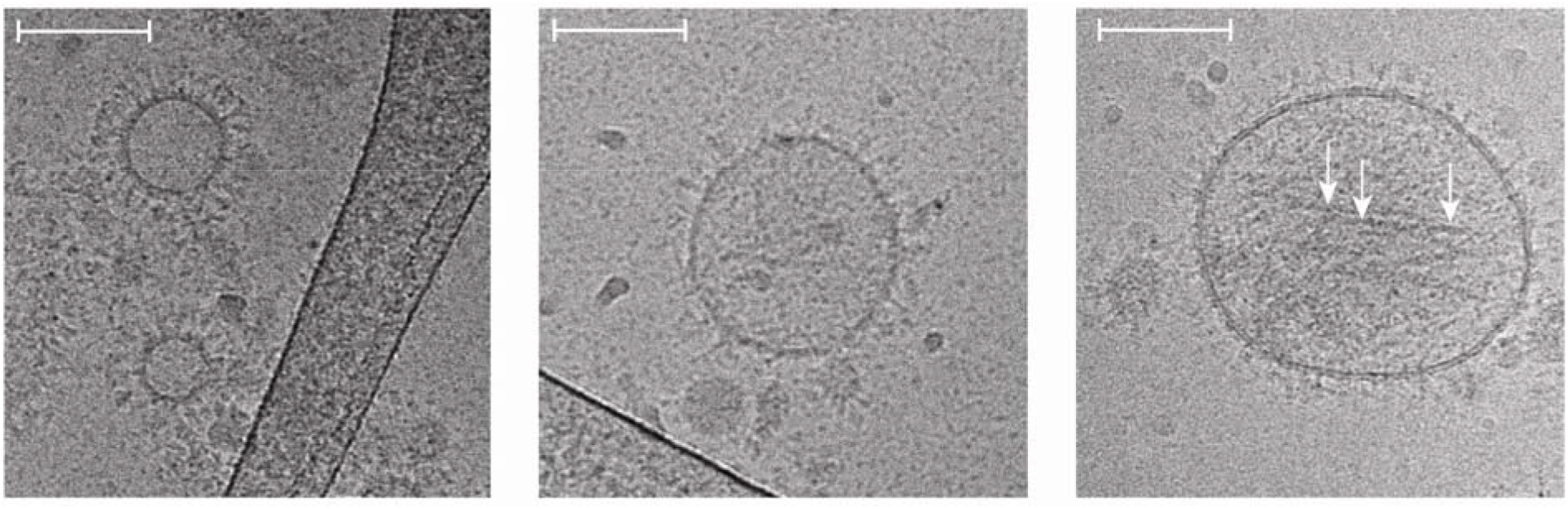
Cryo-electron microscopy images of SARS-CoV-2 VLPs (S^+^ VLPs) The images show different SARS-CoV-2 virus-like-particles (S^+^ VLPs) of approximately 60 to 150 nm in diameter recorded by cryo-electron microscopy (cryo-EM). The particles bear the characteristic corona of radial, dense spike-like proteins protruding from the envelopes’ intact lipid bilayers, which are characteristic for trimers of the viral glycoprotein of coronaviruses, spike (S), as observed for SARS-CoV-2 virions. The particle in the right panel shows elongated structures (white arrows), which might correspond to spike protein protrusions lying down on the vesicle surface, likely caused by surface tension effects prior to rapid freezing of the sample. White scale bar in all three panels corresponds to 100 nm in length.

SARS-CoV-2 virions contain a complex of ribonucleoproteins (N) and the ∼30 kb RNA genome, but S^+^ VLPs seemingly do not contain a similar luminal mass (Fig. 1) probably because the vRNA genome is absent. Together with N, the large vRNA molecule might act as a sizing factor, which could explain the variability in diameter seen in the S^+^ VLP preparations. Other than that, our S^+^ VLPs seem to mimic SARS-CoV-2 virions structurally (Fig. 1).

### Molecular characterization of SARS-CoV-2 VLPs (S^+^ VLPs)

Spike, S, the viral fusion protein (FP) of SARS-CoV-2 is a highly glycosylated type I transmembrane protein which assembles as homotrimers. It encompasses the two domains S1 and S2, which are proteolytically separated by cellular furin protease. After furin cleavage, S1 and S2 remain non-covalently associated (Millet and Whittaker, 2015), but it appears as if furin cleavage is dispensable for infection (Papa et al., 2021).

Because of its central role in viral infection, the correct conformation of S is critical for the tropism and fusogenicity of both SARS-CoV-2 virions and S^+^ VLPs. We used two commercially available antibodies that recognize S1 or S2 together with 43A11, a new, in-house generated high affinity monoclonal antibody (Supplementary Fig. S1), which exclusively recognizes non-dissociated S (but not the single S1 or S2 domains; Fig. 2C) to visualize S1, S2, S full-length (S^FL^) and higher order S^FL^ complexes in S^+^ VLP preparations under reducing and non-reducing conditions. By western blot (WB) analysis (Fig. 2), we confirmed the presence of S1 and S2 domains, S^FL^, trimers of S^FL^ (S^FL^_3_ or S2_3_S1_3_) and other additional S complexes in our S^+^ VLPs preparations. Proteolytic cleavage by furin and possible subsequent dissociation of S1 during viral egress (Papa et al., 2021; Zhang et al., 2020), but also dimeric S^FL^ complexes might generate certain additional higher order complexes (Ou et al., 2020), which we also observed in Fig. 2. In S^+^ VLP preparations from HEK293T cells the majority of S is efficiently cleaved, presumably by furin (Fig. 2B, top panel, reducing conditions), but S1 and S2 domains remain largely complexed in single and higher order S^FL^ conformations (Fig. 2A,C, top panels, non-reducing conditions). The presence of S2, S^FL^ and S^FL^_3_ in lysates of S^+^ VLP producer cells was further confirmed with serum of a COVID-19 convalescent donor (Fig. 2D) and showed a pattern similar to the S2 specific commercial antibody (Fig. 2A, top panel, reducing conditions).

**Figure 2.**
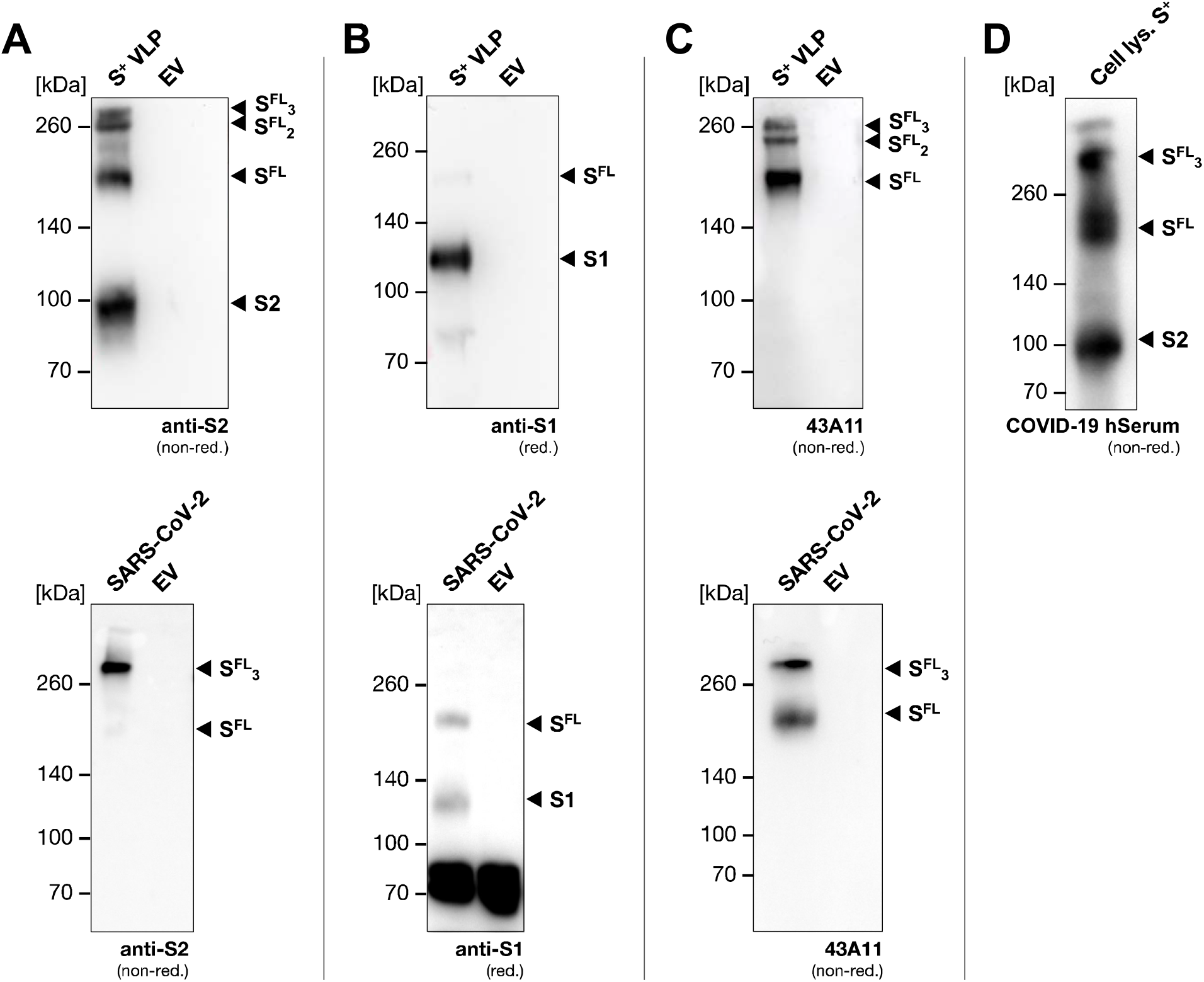
Spike western blot analyses of protein lysates from S^+^ VLPs and SARS-CoV-2 virus stock. Western blot analyses of S^+^ VLPs and extracellular vesicles (EVs) produced in or spontaneously released from HEK293T cells, and SARS-CoV-2 virus stock produced from infected Vero E6 cells are shown. Antibodies are directed against the S1 or S2 domains or recognize the intact, full-length (FL) spike molecule S^FL^. The analyses confirm the presence of spike protein in various states in S^+^ VLPs and SARS-CoV-2 virion preparations but not in EVs which served as negative control. (A, B) Commercially available S2 and S1 specific monoclonal respective polyclonal antibodies detect both spike domains in cell-free preparations of S^+^ VLPs as well as S^FL^ protein (top panels). The S2 domain specific antibody also detects trimeric S^FL^ (S^FL^_3_) and spike complexes of higher order under non-reducing (non-red.) conditions. In SARS-CoV-2 virus stock (bottom panels) the antibodies detect S^FL^ protein and the S1 domain in panels A and B but not the S2 domain. (C) 43A11, an in-house developed, high affinity monoclonal antibody (Supplementary Fig. S1) recognizes full-length spike (S^FL^) exclusively, confirming the presence of mono- and trimeric S^FL^ protein complexes in S^+^ VLPs (top panel) and SARS-CoV-2 virus stock (bottom panel). (D) Serum from a convalescent COVID-19 patient contains antibodies directed against the S2 domain, S^FL^ and S^FL^ multimers in protein lysates from HEK293T cells transiently transfected with expression plasmids encoding the four structural components (S, M, N, E) of SARS-CoV-2.

Parallel to S^+^ VLPs, we analyzed a heat inactivated (56°C, 15 min) SARS-CoV-2 virus stock harvested from infected Vero E6 cells. Compared to S^+^ VLPs, the virus stock showed a similar S composition, but the S2 specific antibody recognized only S^FL^, whereas the S1 antibody detected both S^FL^ and S1. Using the 43A11 monoclonal antibody (mAb), the SARS-CoV-2 virus stock was found to contain S in its distinct trimeric state, but also monomeric S^FL^. Repeated passaging of SARS-CoV-2 on Vero E6 cells can lead to the loss of the furin cleavage site as has been previously reported (Turonova et al., 2020), which might explain the equal fraction of non-cleaved S (Fig. 2B, lower panel) and the absence of the separate S2 domain in the virus stock in Fig. 2A. We conclude that our S^+^ VLP preparations and an authentic SARS-CoV-2 virus stock are similar according to WB analyses but differ with respect to the fraction of furin cleaved S.

We also developed a highly sensitive sandwich enzyme-linked immunosorbent assay (ELISA) to characterize S^+^ VLP and the SARS-CoV-2 virus preparations and to quantify their S content. We used the mAbs 43A11 together with 55E10, a second in-house generated anti-S mAb (Supplementary Fig. S1), which recognize orthogonal, non-overlapping epitopes. As external reference we employed a commercially available recombinant S protein to obtain a calibration curve to assess the amount of S (Fig. 3A). The assay reliably detected S concentrations as low as 3 ng mL^-1^ recombinant protein (Fig. 3A) as well as S protein in S^+^ VLP preparations (Fig. 3B), and was found to be highly specific when probed with control samples consisting of extracellular vesicles (EVs) without a viral fusion protein (Δ vFP EVs). Using this ELISA, we quantified our S^+^ VLP productions and heat inactivated SARS-CoV-2 virus stock (RT-qPCR, ct value of 15.3) and found about 626 ± 20 ng mL^-1^ and 149 ± 6 ng mL^-1^ S protein, respectively, according to the S protein standard (Fig. 3C).

**Figure 3.**
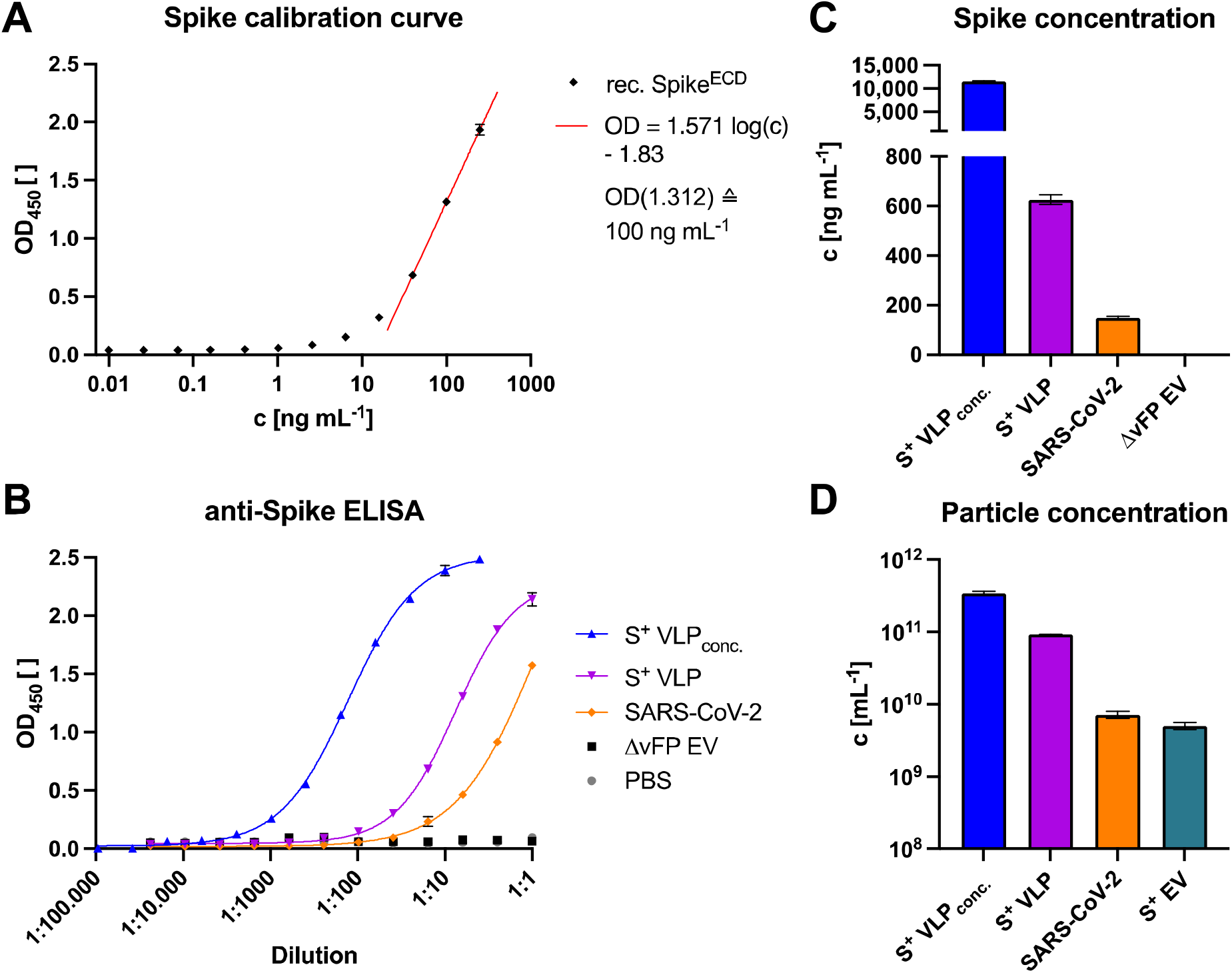
Spike-specific, quantitative sandwich ELISA and nanoparticle tracking analysis (NTA) of S^+^ VLP preparations and heat-inactivated SARS-CoV-2 virus stock. Two in-house generated monoclonal antibodies (43A11 and 55E10), which exclusively recognize two orthogonal, non-overlapping epitopes in S^FL^ protein, were used to establish a sandwich ELISA (enzyme-linked immunosorbent assay) for the quantitation of spike protein in biological samples. (A) Calibration of the spike-specific sandwich ELISA with a commercially available recombinant (rec.) S protein standard, which encompasses the extracellular domain (ECD) of S. The calibration curve of three independent replicates allows for calculating the amount of S protein in samples within the linear range of optical density (OD) values (0.7 ≤ OD ≤ 1.7, r^2^ > 0.99). The detection limit of this assay was estimated to correspond to 3 ng mL^-1^ recombinant S protein. (B, C) Concentrated (conc.) and non-concentrated S^+^ VLPs from supernatants of transiently transfected HEK293T cells were analyzed for their amount of S^FL^ protein. Based on the linear regression function in panel A, the concentration (c) of S was calculated (ng mL^-1^) of three technical replicates in the linear OD range and compared with inactivated SARS-CoV-2 virus stock with a known ct value (15.3) of its vRNA copies according to RT-qPCR. Controls are solvent (PBS) and extracellular vesicles without a viral fusion protein (Δ vFP EVs) harvested from cell culture medium of HEK293T cells transiently transfected with expression plasmids coding for M, N, E and CD63∼HiBiT but omitting S. (D) Nanoparticle tracking analyses (NTA) of three independent preparations of unconcentrated and concentrated S^+^ VLPs (S, M, N, E, CD63∼HiBiT) and S^+^ EVs (S, CD63∼HiBiT omitting M, N and E) from cell culture medium of transiently transfected HEK293T cells are shown. For comparison, NTA data from heat-inactivated SARS-CoV-2 virus stock from infected Vero E6 cells are provided.

### Particle analysis of SARS-CoV-2 VLPs

Next, we quantified the number of total physical particles in our samples via nanoparticle tracking analysis (NTA). Intriguingly, co-expression of M, N, E together with S increased the number of particles drastically compared to transfections of S alone (Fig. 3D), suggesting that SARS-CoV-2 S^+^ VLPs consisting of M, N, E and S evolve via a self-assembling mechanism and egress without the need of non-structural viral proteins, as described for SARS-CoV VLPs (Siu et al., 2008). NTA of our samples indicated that our S^+^ VLP preparations contained 9.1×10^10^ mL^-1^ particles, while comparable preparations of EVs obtained from HEK293T cells after transfection with the S encoding expression plasmid (S^+^ EV in Fig. 3D) only yielded 5.0×10^9^ mL^-1^ particles. The heat inactivated SARS-CoV-2 virus stock was determined to contain 7.2×10^9^ mL^-1^ particles. Total particle counts included 1.5×10^9^ mL^-1^ bovine EVs from fetal bovine serum (FBS) contained in cell culture medium. Bovine EVs corresponded to 2% and 30% of total particle numbers in S^+^ VLP and S^+^ EV preparations, respectively.

To obtain quantitative data of our S^+^ VLPs at the level of single particles, we developed a nano flow technique to assess the fraction of S-positive particles among all particles released by HEK293T cells. Particles were purified from cell culture supernatants and incubated with the dye CellTraceViolet (CTV; Thermo Fisher Sci.), which exhibits fluorescence upon enzymatic ester hydrolysis in the lumen of intact vesicles after membrane penetration. Subsequently, particles were stained with the fluorescently labeled anti-S mAb 43A11 and analyzed by our nano flow technology using a cytometer (CytoFLEX, Beckman Coulter). To distinguish instrument noise from particles we pre-gated on CTV^+^ events and SSC-H (Fig. 4A) and measured the fraction of S-positive particles eventually. S^+^ VLPs, i.e., particles obtained after transient transfection of HEK293T cells with the plasmids encoding S, M, N and E, constituted 37.5% of all CTV^+^ particles. The inactivated SARS-CoV-2 stock contained about 10% S-positive particles (Fig. 4B). Thermal treatment of the SARS-CoV-2 sample might have lowered the esterase activities in both virions and EVs, which might explain the generally lower fraction of CTV^+^ particles (0.11% of all events) in this SARS-CoV-2 virus stock (top panels in Fig. 4B). We therefore estimated the fraction of SARS-CoV-2 virions and S^+^ VLPs to be in a comparable range but S^+^ VLPs were abundant by a factor of three or more.

**Figure 4.**
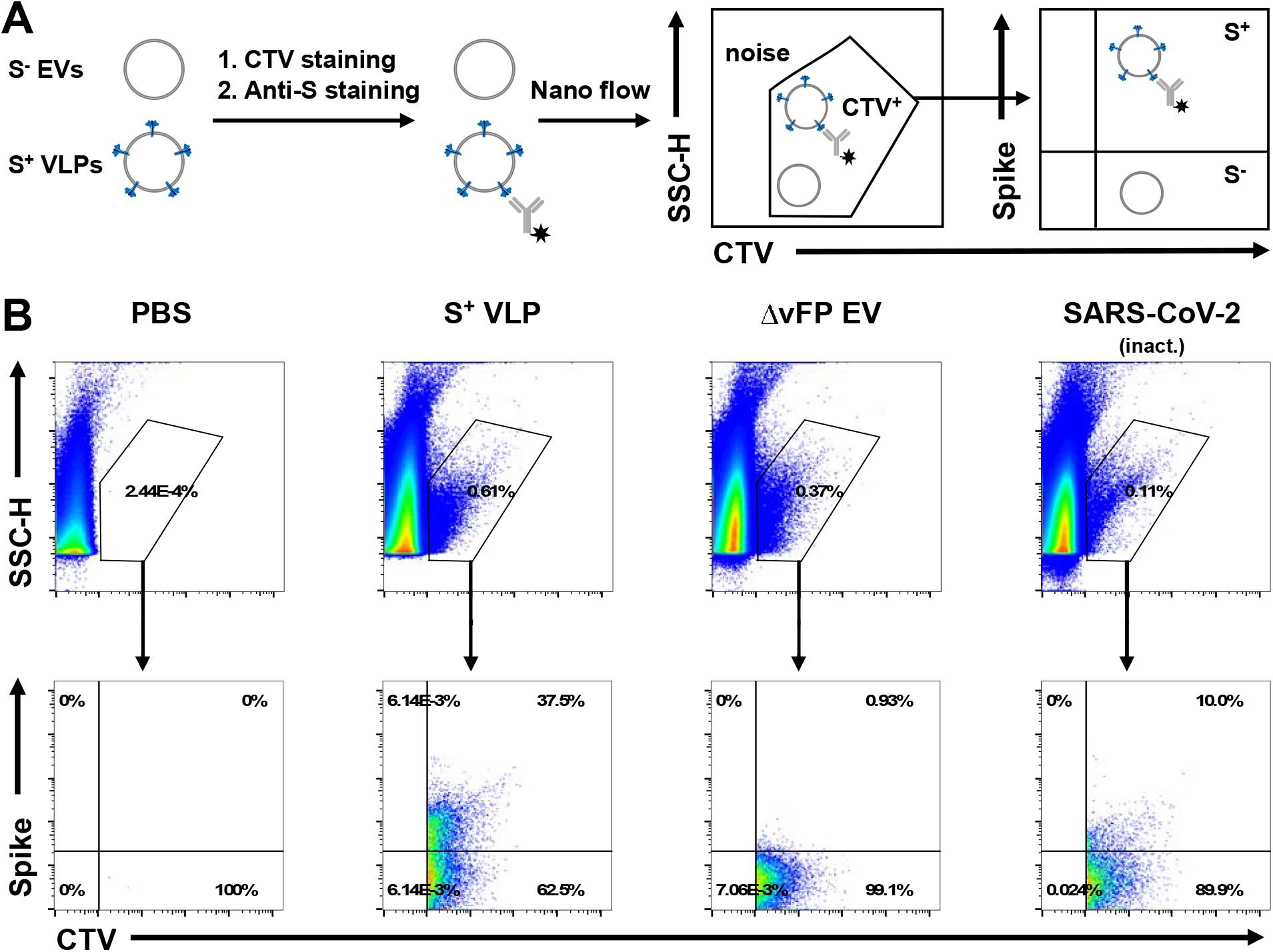
Nano flow technology of S^+^ VLPs and heat-inactivated SARS-CoV-2 virus stock. (A) HEK293T cells transfected with S, M, N, E, CD63∼HiBiT or with M, N, E, CD63∼HiBiT but without S yield S^+^ VLPs or control Δ vFP EVs, respectively. After two rounds of low-speed centrifugation cell culture supernatants containing either S^+^ VLPs or control Δ vFP EVs were stained with the membrane permeable dye CellTraceViolet (CTV), which exhibits fluorescence only upon its uptake followed by esterase activation within the lumen of intact membranous vesicles. A heat-inactivated SARS-CoV-2 virus stock was also stained with CTV for comparison. Subsequently, samples were counter-stained for the presence of surface spike protein using the monoclonal anti-S antibody 43A11. The samples were diluted and analyzed using a CytoFLEX LX flow cytometer (Beckman Coulter Life Sci.). (B) Panels in the top row show all recorded events analyzed for their sideward scatter (SCC-H) using a violet excitation laser (y-axis) and CTV staining (x-axis). CTV^+^ events were gated as shown to identify sub-cellular, intact particles (S^+^ VLPs, Δ vFP EVs, SARS-CoV-2 virus) to distinguish them from instrument noise seen in the PBS control. CTV^+^ events were analyzed for their staining with the anti-S antibody 43A11 coupled to AlexaFluor488 (bottom row of panels). 37.5% S^+^ particles were identified in the preparation of S^+^ VLPs, 10% S^+^ particles were identified in SARS-CoV-2 virus stock and fewer than 1% in control Δ vFP EVs. The low fraction of CTV-positive events in SARS-CoV-2 virus stock (0.11%) compared with preparations of S^+^ VLPs (0.61%) and Δ vFP EVs (0.37%) might be the consequence of a reduced esterase activity in virions (and extracellular vesicles) after heat inactivation at 56°C for 15 min to inactivate viral infectivity.

Our preparations of S^+^ VLPs were found to contain 626 ng mL^-1^ S protein on average (Fig. 3C) and a particle concentration of 9.1×10^10^ mL^-1^ (Fig. 3D), and consisted of 37.5% S^+^ particles (Fig. 4B). Assuming even distribution of S molecules per S^+^ particles and the given molecular weight of 134.4 kDa for the truncated recombinant S protein standard, we calculated a theoretical number of 82 S molecules, corresponding to 27 trimers per S^+^ VLP. This number is in the range described by others for SARS-CoV-2 (Ke et al., 2020; Turonova et al., 2020; Yao et al., 2020). Regarding the inactivated SARS-CoV-2 stock, we determined 149 ng mL^-1^ S protein, 7.2×10^9^ mL^-1^ overall particles and 10% S^+^ particles, which would correspond to 311 S trimers per particle. This surprisingly high number is probably attributable to technical limitations of staining heat inactivated particles along the protocol described above and thus underestimating of the number of S^+^ particles.

### Virus-free neutralization test

Towards a virus-free neutralization test we analyzed the capacity of S^+^ VLPs to fuse with appropriate target cells. First, we transduced various cell lines (HEK293T, LN18, A549, Huh7, Vero and U251MG) to constitutively express human ACE2 and validated its expression by WB. Of these cell lines, Huh7 and Vero cells had previously been used for infection with spike-pseudotyped retrovirus vectors (Yang et al., 2020). Recently, we identified the human U251MG cell line to take up EVs equipped with VSV-G very efficiently suggesting that these cells might also be suitable to act as S^+^ VLP recipient cells (Albanese et al., 2021).

Next, we engineered EVs to contain SARS-CoV-2 spike together with CD63∼BlaM (a chimer of human CD63 and β-lactamase), concentrated them via ultracentrifugation and incubated them with the panel of recipient cells as described above for 4 h. Upon uptake and fusion of these EVs with recipient cells, BlaM translocates to the cytoplasmic compartment of the recipient cells. Only BlaM^+^ cells loaded with an appropriate substrate (CCF4-AM, Thermo Fisher Sci.) convert CCF4-AM to fluorescent CCF4, which is retained in the cytosol and can be quantified via flow cytometry on single cell level as described in the context of HIV-1 (Cavrois et al., 2002), and later by us (Albanese et al., 2021) and others (Carlon- Andres and Padilla-Parra, 2020; Cavrois et al., 2014; Desai et al., 2017; Feeley et al., 2011; Jones and Padilla-Parra, 2016).

ACE2^+^ Vero cells took up S^+^ EVs but not control EVs lacking a viral fusion protein (Δ vFP EVs) (Supplementary Fig. S2A). Yet only about 21% of Vero cells turned BlaM-positive even with a high dose of S^+^ EVs, while 75% of all cells became positive with control EVs equipped with CD63∼BlaM and VSV-G (vesicular stomatitis virus glycoprotein G) as a viral fusion protein (VSV-G^+^ EVs; Supplementary Fig. S2B). In contrast, up to 97% ACE2^+^ U251MG cells became BlaM-positive with S^+^ EVs, while cells incubated with Δ vFP EVs remained BlaM negative (Supplementary Fig. S2C, D). The fusion of ACE2^+^ U251MG cells with S^+^ or VSV-G^+^ EVs was equally efficient in this setting (Supplementary Fig. S2C, D). To demonstrate the endosomal uptake and fusion of EVs with the recipient cells, we pre-treated ACE2^+^ U251MG cells with chloroquine prior to incubation with S^+^ EVs (Supplementary Fig. S2E). Chloroquine deacidifies endosomes and thus also inactivates the endosomal-pH-dependent cysteine protease Cathepsin L (CTSL), which primes S for SARS-CoV-2 entry in certain cell lines *in vitro* (Hoffmann et al., 2020a; Hoffmann et al., 2020b). Thus, delivery of BlaM was found to be strictly dependent on endosomal processing of S in recipient cells.

Next, we used ACE2^+^ U251MG cells in preliminary neutralization experiments to test and quantify the reduction of S-mediated fusion of BlaM equipped S^+^ EVs by neutralizing serum antibodies. Sera from SARS-CoV-2 vaccinees and COVID-19 convalescent patients displayed dose-dependent neutralization, while sera from healthy and naïve donors barely showed any effect (Supplementary Fig. S2E). We concluded that all steps of the S-mediated and ACE2-dependent cellular uptake of S^+^ EVs are highly reminiscent of SARS-CoV-2 infection.

### Optimization of the VLP fusion assay

The applicability of the assay described above is limited as its readout relies on flow cytometry, requires an overnight incubation step for the intracellular accumulation of CCF4 and depends on concentrated S^+^ EVs preparations. We therefore developed this assay further by replacing BlaM with nano-Luciferase (nLuc). To avoid known background problems due to protein leakage of intact luminescent nLuc, we adapted its split variant consisting of an incomplete and inactive nLuc polypeptide (LgBiT) and the self-associating activator peptide of 11 amino acids (HiBiT) for our purposes (Dixon et al., 2016; Miyakawa et al., 2020; Yamamoto et al., 2019).

Similar to BlaM, we fused HiBiT to the C terminus of CD63 (CD63∼HiBiT) (Somiya and Kuroda, 2021) to be incorporated into S^+^ particles. As recipient cells, we engineered ACE2^+^ U251MG cells to constitutively express *N*-myristoylated LgBiT (NM∼LgBiT) as membrane-associated reporter enzyme. This system proved to be almost free of leakage as HiBiT and LgBiT are tightly associated with the cellular and EV membranes, respectively, and are not secreted in detectable amounts. In the split nLuc system, the turnover of a suitable substrate is only catalyzed upon successful intracellular reconstitution of both parts of nLuc (Dixon et al., 2016). As a consequence, this assay has a very low background, which otherwise is a major problem when working with fully active enzymes as protein reporters.

We also turned from S^+^ EVs to SARS-CoV-2 VLPs, i.e., S^+^ VLPs encompassing all four structural SARS-CoV-2 proteins, because co-expression of M, N and E together with S and CD63∼HiBiT (Fig. 5A) led to higher particle numbers and enhanced fusogenicity of VLPs with ACE2^+^ U251MG cells (Fig. 3D). We determined the optimal stoichiometry of plasmid DNAs in this practical, readout-based approach to obtain S^+^ VLPs which resemble SARS-CoV-2 in many aspects as documented by cryo-EM, WB, ELISA, NTA and nano flow technology as shown in Fig. 1 to 4. The engineered S^+^ VLPs enter ACE2^+^ recipient cells in a receptor-dependent manner, require correct processing of S by proteases and escape from endosomes via the post-fusion conformation of S, very reminiscent of infectious SARS-CoV-2 virions. Upon endosomal fusion the membrane anchored HiBiT is delivered into the cytoplasm of the LgBiT^+^ recipient cell, where the functional enzyme reconstitutes *in situ* to support substrate turnover and emission of light as shown schematically in Fig. 5C. A scientific animation demonstrates this simple but very efficient principle (https://youtu.be/6wckXobT_bM).

**Figure 5.**
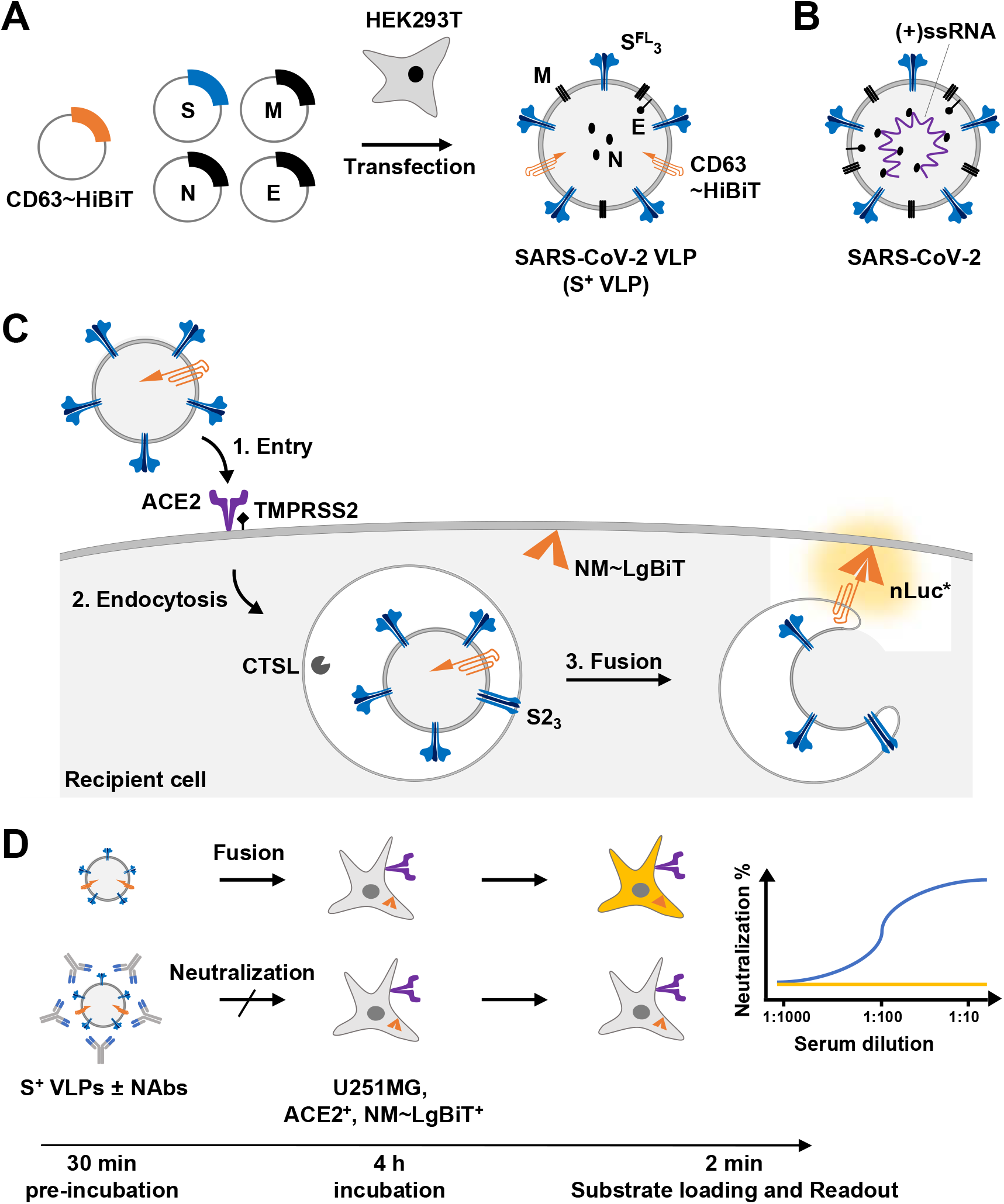
VLP neutralization test (VLPNT) (A) Engineered VLPs were generated *in vitro* by transient co-transfection of HEK293T cells with an optimized ratio of expression plasmids encoding the four SARS-CoV-2 structural proteins S, M, N, E and a chimeric membrane anchored activator peptide (CD63∼HiBiT). The resulting particles were termed S^+^ VLPs and obtained from conditioned cell culture medium three days after DNA transfection. (B) Schematic view of a SARS-CoV-2 virion with the four structural proteins S, M, N and E and the viral genome of positive sense, single-stranded RNA complexed with N. (C) Basic steps of VLP entry and reconstitution of nano Luciferase (nLuc). Similar to infection with SARS-CoV-2, spike, the trimeric viral fusion protein in the envelope of S^+^ VLPs (Fig. 1) mediates attachment to the host cell receptor ACE2, triggering endocytosis and proteolytic processing by TMPRSS2 or CTSL. The ensuing fusion of the S^+^ VLP envelope with cellular membranes exposes the HiBiT activator peptide to make contact with *N*-myristoylated LgBiT (NM∼LgBiT), which is stably expressed in the cytoplasm of the ACE2^+^ target cell. Upon *in situ* reconstitution of the functional nano Luciferase (nLuc*) reporter addition of substrate will induce bioluminescence, which can be quantified in a standard luminometer in 96-or 364-well cluster plates. (D) To test body fluids for the content of neutralizing SARS-CoV-2 antibodies (NAbs), S^+^ VLPs are pre-incubated with serial dilutions (on the basis of 2) of the samples for 30 min. Suitable medical samples are sera of COVID-19 patients, vaccinated or naïve individuals or other body fluids such as saliva or nasal excretions. SARS-CoV-2 NAbs will interfere with all steps of S^+^ VLP attachment to ACE2, receptor-mediated intake, endosomal fusion of the VLP envelope with the endosome and escape to the cytoplasm. Target cells are U251MG cells engineered to express both ACE2 and NM∼LgBiT (LgBiT). Upon encounter with S^+^ VLP-borne CD63∼HiBiT, NM∼LgBiT is reconstituted into a fully functional nLuc reporter enzyme, which can be quantitated. Neutralizing SARS-CoV-2 antibodies reduce or even block the delivery of the CD63∼HiBiT activator entirely, which can be quantified in a standard clinical laboratory with aid of a luminometer and within 4.5 hours. A freely accessible scientific animation narrates the principle of the VLPNT (https://youtu.be/6wckXobT_bM).

Analogous to the EV system with BlaM as reporter protein, we assessed whether S^+^ VLPs fuse exclusively with susceptible ACE2^+^ cells via the viral entry factor S. We generated S^+^ VLPs (S, M, N, E, CD63∼HiBiT)^+^, VSV-G^+^ EVs (VSV-G, CD63∼HiBiT)^+^ and Δ vFP EVs (M, N, E, CD63∼HiBiT)^+^, incubated them with ACE2^+^ or ACE2^-^ NM∼LgBiT^+^ U251MG cells for 4 h and quantified their fusion with the different target cells. As expected, S^+^ VLPs fused exclusively with ACE2^+^ cells but not with ACE2^-^ cells (t-test, p<0.0001) (Fig. 6). Furthermore, fusion relied strictly on the presence of a viral fusion protein such as S because Δ vFP EVs were barely taken up (t-test, p<0.0001). In contrast, VSV-G^+^ EVs fused with ACE2^+^ and ACE2^-^ cells at similar levels due to VSV-G’s broad tropism (t-test, p value non-significant) (Fig. 6). We conclude that this system meets all requirements of a S^+^ VLP-based neutralization test.

**Figure 6.**
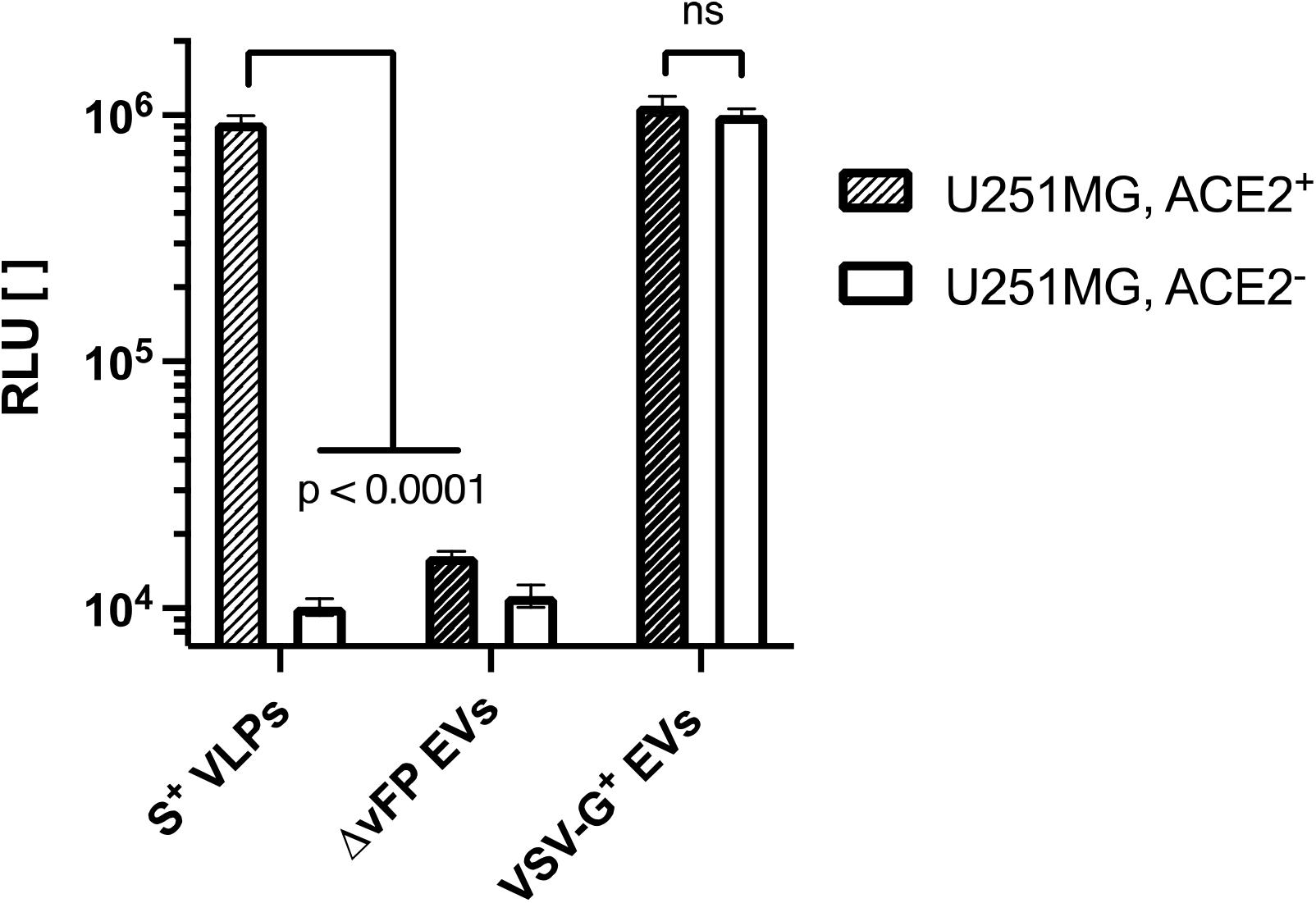
Specificity and tropism of S^+^ VLPs. U251MG cells were transfected to express NM∼LgBiT constitutively. In addition, we established a derivative of these NM∼LgBiT cells with the human ACE2 receptor to obtain both ACE2-negative (ACE2^-^) and positive (ACE2^+^) target cells. The two cell lines were incubated with S^+^ VLPs carrying CD63∼HiBiT, with EVs without a viral fusion protein (ΔvFP EVs) obtained from supernatants of HEK293T cells after transient transfection of expression plasmids encoding M, N, E and CD63∼HiBiT (but not S) or with EVs from HEK293T cells after transient transfection of expression plasmids encoding CD63∼HiBiT and protein G of the vesicular stomatitis virus (VSV-G^+^ EVs). The specificity of spike-mediated, ACE2-dependent fusion of all three particle classes was validated measuring luciferase activities upon reconstitution of the split nano-luciferase in the indicated cell types. The results document the high specificity of S^+^ VLPs to fuse exclusively with ACE2^+^ cells, while Δ vFP EVs yielded background signals, only. Both cell lines were equally positive when incubated with control VSV-G^+^ EVs indicating their ACE2 receptor independent uptake and fusion, reflecting the broad tropism of VSV. Data are based on at least four independent experiments. p-values of three independent t-tests are indicated (ns; not significant).

### VLP neutralization test (VLPNT)

S-specific NAbs can prevent entry of SARS-CoV-2 virions into host cells and thus protect from viral infection via diverse modes of actions. Most antibodies block the attachment of S to ACE2 receptors by binding to the receptor binding motif (RBM) of S1 (Niu et al., 2021; Piccoli et al., 2020) or stall S in its closed, i.e., receptor binding domain (RBD) “down” pre-fusion conformation (Tortorici et al., 2020). Yet, certain S-specific NAbs also neutralize without disrupting the ACE2 interaction (Brouwer et al., 2020). Possible other mechanisms include the inhibition of proteolytic processing of S by, e.g., TMPRSS2 or CTSL or the interference with the heptad repeats or glycosylated surfaces in S2, which are required to promote the fusion of the viral envelope with the endosomal membrane as described in the context of SARS-CoV or MERS-CoV (Keng et al., 2005; Shanmugaraj et al., 2020). NAbs with their multiple mechanisms to disrupt S functions also reduce, interfere or even block S^+^ VLPs and the delivery of CD63∼HiBiT to susceptible target cells. As a result, reduction of luminescence from reconstituted nLuc might likely correlate with SARS-CoV-2 neutralization (Fig. 5D).

Towards a VLP-based virus neutralization test (VLPNT), we incubated a defined amount of S^+^ VLPs with serial dilutions (starting from 1:10 to 1:1,800) of sera from acute or convalescent COVID-19 patients, COVID-19 vaccinees or healthy, naïve donors and quantified the resulting dose-dependent neutralization as shown in Fig. 7A. Mean luminescence level of S^+^ VLPs, only, was set to 0% neutralization; while background luminescence obtained with Δ vFP EVs was set to 100% neutralization. The half maximal neutralization titer was determined by extrapolating sigmoidal curve values that correspond to 50% signal reduction after background correction. This value was termed VLPN_50_. It is considered equivalent to VNT_50_ and PRNT_50_ of conventional VNTs (cVNTs) and plaque reduction neutralization tests (PRNTs), respectively.

**Figure 7.**
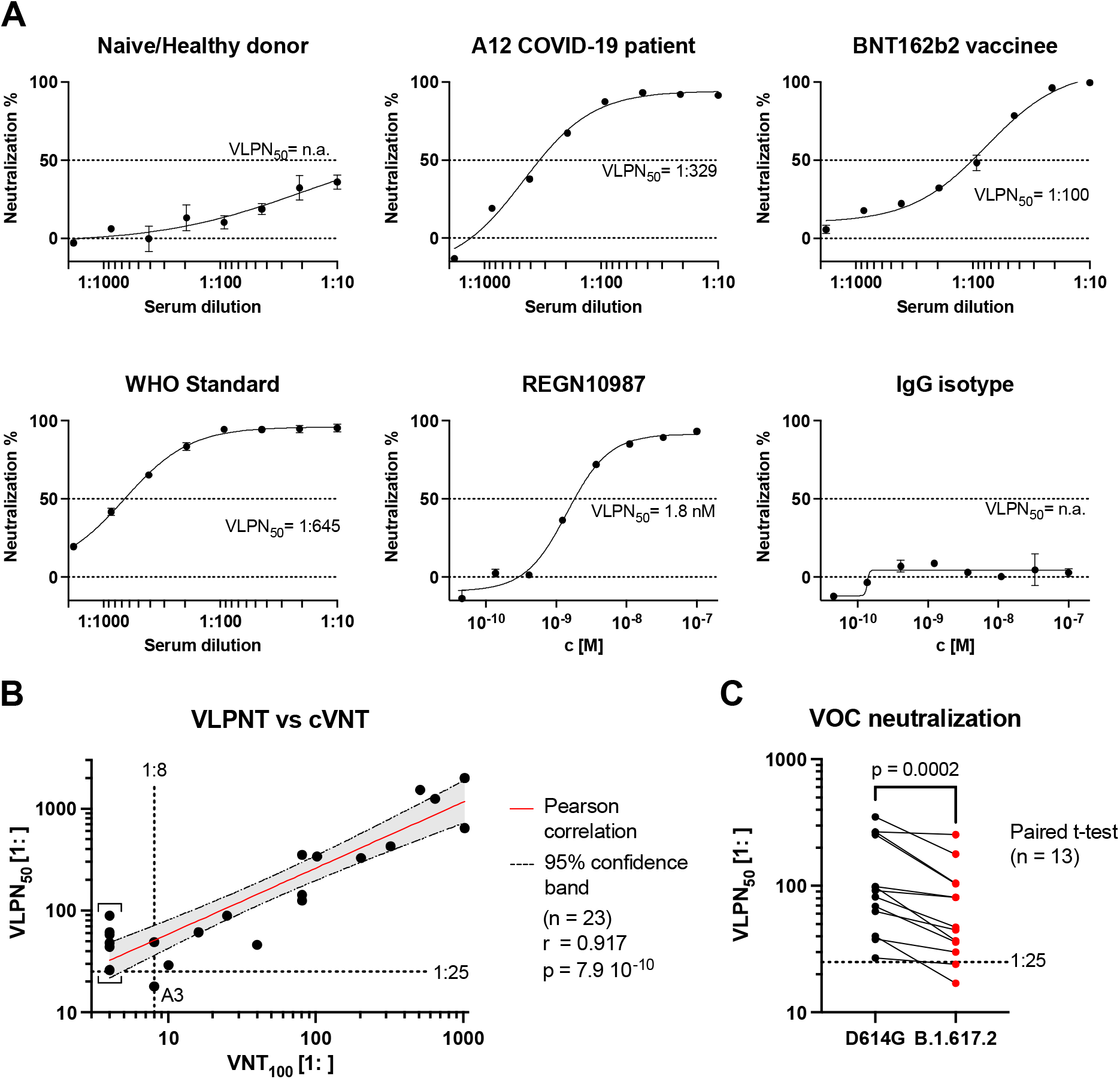
Correlation of VLPNT and cVNT data and VOC cross-neutralization using sera from COVID-19 patients and vaccinees. (A) VLPNT neutralization data with dilutions of sera obtained from three individuals (a naïve healthy donor, the COVID-19 patient A12 and a BNT162b2 vaccinee) and the WHO reference serum (NIBSC 20/136) as international standard are shown. The graphs are examples and include mean neutralization results from three independent biological replicates. Serum dilution which resulted in half maximal signal reduction, equivalent to 50% neutralization, was termed VLPN_50_ titer. A recombinant human mAb, REGN10987 with known neutralizing activity showed potent neutralization in the assay at low molar concentration (c), while an IgG isotype control mAb did not show any neutralizing activity. (B) Validation of the VLPNT by correlating VLPN_50_ titers versus VNT_100_ titers obtained in a cVNT, the gold standard using infectious SARS-CoV-2 virus. Pearson correlation data (sample size n, coefficient r and p-value) of 23 sera from confirmed COVID-19 patients are shown and the linear relationship is indicated. The sample denoted A3 is a single serum sample referred to in the Result section. Results below the dotted horizontal line denote sera which scored negative in the VLPNT. Results left of the dotted vertical line denote sera which scored below the limit of detection (1:8) in the cVNT; these VNT_100_ values were defined as 1:4 and indicated by square brackets. (C) VLPNT with a SARS-CoV-2 variant of concern (VOC) compared with Wuhan-2019 D614G. S^+^ VLPs were harvested from supernatants of HEK293T cells transiently transfected with expression plasmids encoding either Wuhan-2019 D614G or Delta mutant B.1.617.2 S protein together with M, N, E, and CD63∼HiBiT. NAbs in sera of 13 COVID-19 vaccinees were cross-neutralizing, but less potent in neutralizing the δ-mutant B.1.617.2 compared with the Wuhan-2019 D614G mutant. Data were statistically analyzed using the paired t-test.

Sera of naïve donors resulted in no to little signal reduction, i.e., weak neutralization even at high serum concentrations. Of 12 sera from healthy, naïve donors obtained from mid 2019 and earlier, most did not reach 50% neutralization and a few showed VLPN_50_ titers of 1:16 or lower (Supplementary Fig. S3A). Sera of 13 COVID-19 vaccinees after prime-boost immunization from 2021 showed high neutralization potential consistently up to 100% inhibition of S^+^ VLPs fusion and VLPN_50_ titers of 1:27 to 1:352 (Supplementary Fig. S3A). Data on individual titers and types of vaccines which the donors received can be found in Supplementary Fig. S3D and Supplementary Tab. S1. Based on the maximal titers of naïve sera in our VLPNT, we added an additional margin and defined a preliminary minimal cutoff of 1: ≥25 to classify samples with neutralizing activities.

As there is considerable demand for global harmonization and standardization of NAb titers obtained from different laboratories using different versions of SARS-CoV-2 VNTs (Kristiansen et al., 2021), the World Health Organization (WHO) supplies laboratories with a standard plasma pooled from 11 British patients, who recovered from COVID-19 (Mattiuzzo *et al*., 2020, WHO/BS/2020.2403). We applied this standard (NIBSC 20/136) with a defined neutralization activity of 1000 International Units per mL (IU mL^-1^). VLPN_50_ values of multiple individual experiments with the WHO standard were obtained (Supplementary Tab. S1) and used to convert our serum VLPN_50_ titers to harmonized titers expressed as IU mL^-1^ as shown in Supplementary Fig. S3A, right panel. In six independent VLPNT experiment with the WHO standard, we calculated a coefficient of variation (CV) for log_10_ transformed titers of CV = 0.04 and CV = 0.23 for untransformed titers, for the within-laboratory repeatability and the resulting imprecision of quantification, respectively. Clearly, additional experiments and standardizations have to be conducted to generate data for an assessment according to the Clinical Laboratory Standards Institute (CLSI) guidelines (Chesher, 2008).

We also used our VLPNT to test two potent neutralizing recombinant mAbs, REGN10987, imdevimab and REGN10933, casirivimab, which have been approved under emergency use authorization for the treatment of COVID-19 by the Food and Drug Administration (FDA) (2020; Baum et al., 2020; DeFrancesco, 2020). Their VLPN_50_ were found to be 1.8 nM and 7.3 nM, respectively, while a human IgG isotype control mAb did not show any neutralization (Fig. 7A).

Based on these results we concluded that the VLPNT qualifies for the measurement of neutralizing activities in clinical samples.

### VLPNT and cVNT compared

For the validation of our VLPNT we compared the VLPN_50_ titers of a defined set of sera with results from a cVNT using infectious SARS-CoV-2, the ‘gold standard’ in the field (von Rhein et al., 2021). Towards this end, we tested 23 well documented serum samples from patients with confirmed SARS-CoV-2 infection under double-blinded, randomized conditions in our VLPNT. Relevant details about their clinical status, PCR results, SARS-CoV-2 strains and neutralizing serum titers data had been collected prior to our analyses (Supplementary Tab. S1).

When analyzed in our test, serum VLPN_50_ titers of the 23 COVID-19 sera varied from 1:18 to 1: >2,000 and therefore scattered wider than serum titers from vaccinees, even though the medians were similar (Supplementary Fig. S3A, left panel). Data on the individual titers and the patients’ clinical symptoms can be found in Supplementary Fig. S3C and Supplementary Tab. S1. Due to the previously defined cutoff for the VLPN_50_ of 1: ≥25 (Supplementary Fig. S3A, left panel), a single COVID-19 sample (sample A3) was classified as non-neutralizing even though the highest titer of a naïve, SARS-CoV-2 negative serum was found to be 1:16 (Supplementary Fig. S3A).

The 23 COVID-19 samples were also tested in a cVNT with replication competent SARS-CoV-2 virus. The titers were determined based on 100% reduction of CPE (VNT_100_) and their values ranged from 1:8 to 1: >1024 (Fig. 7B). In the cVNT, 17 out of 23 samples were found to have neutralizing activity with VNT_100_ 1: ≥8, while the 6 remaining sera performed below the limit of detection (LOD) and were set to 1:4 per definition. A comparison of the 23 COVID-19 samples in both tests found 16 sera to be concordant positive (CP), none was concordant negative (CN), whereas 7 sera were found to be discrepant (D) (Supplementary Fig. S3B, left panel). This discrepancy likely resulted from experimental differences between the two tests (Khoury et al., 2020). The cVNT ‘gold standard’ relies on 100% reduction of CPE (VNT_100_) while the VLPNT scores at 50% reduction of S^+^ VLP fusion (VLPN_50_) suggesting that the latter test might be more sensitive to identify serum samples with weakly neutralizing activities.

Excluding the 6 sera tested in the cVNT with VNT_100_ values below LOD (samples in brackets in Fig. 7B), we analyzed the performance of the VLPNT using the 17 COVID-19 samples which scored positive in the cVNT along with 12 sera from healthy, naïve donors from mid 2019 and earlier as controls (Supplementary Fig. S3B, right panel). Of the 17 COVID-19 samples, the VLPNT classified one sample to be false negative, while all 12 healthy naïve sera were considered true negative. Therefore, the sensitivity of our VLPNT was calculated to be 94%, while its specificity was 100% (n = 29). Based on the small sample number the positive predictive value (PPV) was calculated to be 100% suggesting that our SARS-CoV-2 VLPNT has the potential to become a reliable diagnostic tool.

We also compared the VLPNT with the cVNT by correlating the VLPN_50_ titer results of all 23 COVID-19 samples with the respective VNT_100_ titers, regardless of whether they were considered CP, CN or D (Fig. 7B). The Pearson coefficient of log_10_ transformed data (n = 23) revealed a highly significant positive correlation (r = 0.917 and p = 7.9×10^−10^). Given the very strong correlation, the VLPNT is not only a qualitative test for the *in vitro* diagnostics of NAbs but also yields reliable titers of NAbs faithfully reflecting titers obtained in a cVNT with infectious SARS-CoV-2.

### Test adaptation to Delta-SARS-CoV-2 (B.1.617.2)

Finally, we adapted the VLPNT to test and compare the currently predominant variant of concern (VOC) of SARS-CoV-2 by mutating S (D614G) to S (B.1.617.2, δ-or Delta variant) and generated S^+^ (B.1.617.2) VLPs. We re-tested all 13 sera of SARS-CoV-2 vaccinees (Supplementary Fig. S3D) in the VLPNT with S^+^ (B.1.617.2) VLPs, determined their VLPN_50_ titers and compared them to titers obtained with the D614G S variant (Fig. 7C). Pearson coefficient of log_10_ transformed data (n = 13) was calculated and showed strong positive correlation (r = 0.911; p = 1.5×10^−5^). Apparently, NAbs induced by different SARS-CoV-2 vaccines based on Wuhan-2019 spike cross-neutralized VOC B.1.617.2, but neutralization titers were significantly lower as shown by a paired t-test of log_10_ transformed data (p = 0.0002, n = 13). While the mean titer for D614G S^+^ VLPs was found to be 1:135, the mean titer of 1:80 obtained with B.1.617.2 S^+^ VLPs was reduced by a factor of 0.59. A reduced capacity of convalescent or vaccinee sera to cross-neutralize B.1.617.2 has been demonstrated previously in a pVNT (Liu et al., 2021a), but also in a cVNT (Planas et al., 2021). Thus, our finding appears to be consistent with published work. The successful adaptation of the VLPNT to a prevalent VOC indicates that our test is flexible and versatile to address and answer questions on cross-neutralizing antibodies between existing and future SARS-CoV-2 variants of concern including the currently spreading B.1.1.529 VOC, named Omicron.

## Discussion

Serum antibody levels are a predictive and easily accessible medical parameter of individual immune protection from viral infections and related diseases. For SARS-CoV-2, especially NAb titers were shown to correlate with clinical protection from COVID-19 (Khoury et al., 2021). In clinical virology, the detection and quantitation of NAbs in biological samples is commonly performed with conventional VNTs (cVNTs), which mostly rely on *in vitro* cytopathic effects (CPE) and the formation of plaques in infected cell monolayers. NAbs interfere with cellular infection but the formation of plaques depends on replication competent virus. As a consequence, these tests require appropriate containment and regulatory measures. cVNTs with SARS-CoV-2 for instance require a BSL-3 containment. Moreover, the tests take several days and they are difficult to normalize and to standardize between different laboratories. Plaque formation requires visual microscopic inspection by trained personnel and PRNTs are thus labor intensive and cumbersome. Therefore, surrogate methods have been invented and used, which are commonly based on pseudotyped retro-or lentiviruses so-called pVNTs. The tests, which are manageable in BSL-2 laboratories rely on *de novo* expression of a reporter enzyme or fluorescent protein encoded by the viral vector, but they still require up to three days for final assessment.

The artificial, replication deficient retro-or lentiviral vectors can be problematic because viral glycoproteins, which mediate attachment, uptake and fusion with cellular membranes might perform differently in the context of viral vectors compared to their authentic viral host (Khoury et al., 2020; Syed et al., 2021). S-pseudotyped retro-or lentiviral vectors mostly bud from the plasma membrane where S is incorporated into the envelope of the vector with uncertain stoichiometry (Giroglou et al., 2004; Syed et al., 2021). Very much in contrast, the envelopes of coronaviruses carry a discrete number of S trimers per particle, which assemble in the ER-Golgi intermediate compartment together with M, E and N in a given, optimal stoichiometry (Ujike et al., 2016).

We performed such pVNT for SARS-CoV-2, produced S-pseudotyped, replication incompetent MLV retroviruses, pre-incubated them with different human sera, infected hACE2^+^ Vero cells, cultivated them for 48 h to allow expression of the GFP reporter gene and analyzed them by flow cytometry and fluorescence microscopy (Supplementary Fig. S4). On average about 80% of cells were infected, while pre-incubation of the test vectors with sera from COVID-19 convalescent patients and COVID-19 vaccinated individuals reduced the fraction of infected cells in a dose dependent manner. Conversely, sera from healthy naïve individuals did not show an effect. pVNTs allow to quantitate NAbs with SARS-CoV-2 specificity, but they are biologically distant from infectious SARS-CoV-2 stock, they take several days and they depend on qualified S-pseudotyped virus stocks with reproducible vector content and infectivity.

Miyakawa et al. (Miyakawa et al., 2020) considered the unfavorable time-to-readout problem of pVNTs and replaced the HIV-borne reporter gene with a reporter protein consisting of the HIV capsid protein with a carboxy-terminal HiBiT domain following the principle proposed by Cavrois et al. in 2002 (Cavrois et al., 2002). Similar to our approach (Fig. 5) recipient Vero cells expressed LgBiT constitutively to monitor the fusion of the S-pseudotyped HIV particles, which shortens the time-to-readout to 3 to 4 hours (Miyakawa et al., 2020). This approach, however, does not solve or address the relatively weak correlation of pVNTs and live-virus cVNTs observed in clinical samples (Feng et al., 2021), which probably reflects differences in viral morphogenesis, egress, composition of S-pseudotyped retro- and lentiviral vector particles and their mode of uptake by recipient cells compared to SARS-CoV-2 virions.

For SARS-CoV (Hsieh et al., 2005), and later SARS-CoV-2, VLPs were shown to mediate transport and delivery of reporter transcripts that encompass cis-acting packaging signal sequences mimicking the generation of authentic SARS-CoV-2 and their infectivity (Syed et al., 2021). This is a major achievement, but the readout still depends on *de novo* expression of the reporter transcript encoding luciferase in this report.

Our test overcomes the current limitations of quantitating SARS-CoV-2 NAbs as it is based on harmless, non-infectious VLPs engineered to be authentic morphological and functional mimics of SARS-CoV-2 virions. Firstly, we optimized conditions for *in vitro* generation of such S^+^ VLPs and characterized them together with a sample of heat-inactivated SARS-CoV-2 stock using standard approaches together with a novel nano flow technology. By cryo-EM, we confirmed the presence of intact, native S^+^ VLPs with the characteristics of coronaviruses in our preparations. Furthermore, the S^+^ VLPs contain S^FL^ trimers, a molecular concentration of S very similar to authentic SARS-CoV-2 virions and particle number and ratio of S^+^ particles comparable to virus stocks. We calculated the number of S trimers per particle to be 27 and therefore well within the range of published figures which vary from 24 to 40 (Ke et al., 2020; Turonova et al., 2020; Yao et al., 2020).

Secondly, we engineered S^+^ VLPs to carry a chimeric, membrane anchored and luminally oriented enzyme (BlaM) or an activator peptide (HiBiT) and generated hACE2^+^ Vero cells and a hACE2^+^ U251MG cell line, which carries hACE2^+^ together with an inactive, membrane-associated split reporter nLuc enzyme (LgBiT). In quantitative fusion experiments with S^+^ VLPs we showed that only ACE2^+^ cells take up S^+^ VLPs, while ACE2^-^ cells were not susceptible as expected. Uptake strictly depended on spike, the viral entry factor and one of the three SARS-CoV-2 membrane proteins, indicating that our S^+^ VLPs do not only structurally and molecularly resemble SARS-CoV-2 virions but also share their specific fusogenic characteristic and tropism. A limitation of this *in vitro* cell model is that CTSL but not TMPRSS2 contributed to spike cleavage and processing unlike the infection pathway in human lung cells. cVNTs that use African green monkey Vero cells as targets for SARS-CoV-2 also suffer from this limitation, which can be easily mitigated by modifying the target cells to express hTMPRSS2 but not CTSL.

With the thoroughly characterized S^+^ VLPs and our reporter system, we went on and demonstrated the proof-of-concept for a VLPNT. We incubated S^+^ VLPs with 23 sera from COVID-19 and convalescent patients, 13 COVID-19 vaccinees and two neutralizing recombinant mAbs (REGN10987, imdevimab and REGN10933, casirivimab) and quantitated the resulting reduction of VLP-cell fusion. We controlled our test with 12 sera from healthy and naïve donors and an IgG isotype mAb, for which we did not find significant neutralization capacities. We quantitatively evaluated all samples to determine their individual titers. Based on the tested cohort of donors we defined a titer of 1: ≥25, to classify a sample to contain SARS-CoV-2 specific neutralizing antibodies (NAbs). Below this level matrix effects from healthy, naïve sera came into play, which is why even weaker neutralizations cannot be distinguished from artefacts. For the 23 COVID-19 samples we correlated the titers with the double-blinded results from a cVNT with SARS-CoV-2, resulting in convincing quantitative concordance. Thereby, we verified our VLPNT versus a cVNT and found good preliminary sensitivity and specificity of all test parameters.

Determining NAb levels is of epidemiological and clinical relevancy, as a reduction below a certain threshold can impact protection from infection and increase the personal risk to develop COVID-19. Khoury *et al*. found that 20% of the mean NAb level detected in convalescent sera provided 50% protection from symptomatic disease (Khoury et al., 2021). Based on these data, a minimal NAb titer could be specified, below which booster shots might become advisable.

Since the start of the pandemic in December 2019, the SARS-CoV-2 genome has undergone several mutations especially in the S protein. The original Wuhan-Hu-1 isolate (Wu et al., 2020) rapidly gained the D614G spike mutation (Korber et al., 2020) and evolved further to the B.1.617.2 VOC, which is the predominant variant in USA and Europe as of November 2021 (data of cdc.gov and ecdc.eu). In our VLPNT, emerging VOCs such as the very recently identified B.1.1.529 Omicron VOC can be tested as to whether they present as immune escape mutants and resist neutralization when using sera from vaccinees or convalescent patients vaccinated resp. infected with previous SARS-CoV-2 strains. This is because S^+^ VLPs that contain newly identified spike variants can be produced and validated rapidly and with ease using the WHO reference standard. Along this line, we re-analyzed sera from 13 vaccinees using S^+^ VLPs with B.1.617.2 spike and found cross-neutralization but also a significant reduction of neutralization titers concordant with published findings (Liu et al., 2021a; Planas et al., 2021). In principle, our VLPNT should also be capable of identifying antibody dependent enhancement effects (ADE), which are known for other viruses and has also been reported in the context of SARS-CoV-2 infection (Liu et al., 2021b).

In summary, we present a rapid and safe virus-free yet authentic test to quantitate SARS-CoV-2 NAbs. The structurally and biochemically well characterized S^+^ VLPs faithfully replicate the initial steps of SARS-CoV-2 infection and allow their quantitative analysis. The VLPNT correlates very well when put to the test with a set of COVID-19 patient sera analyzed in a benchmark cVNT with fully infectious, calibrated SARS-CoV-2 stock. The technology fulfills important automatization and upscaling criteria to be suitable for high-throughput screening approaches in low containment laboratories for potential neutralizing mAbs or anti-viral drugs. In principle, this assay is adaptable to other (emerging) enveloped viruses such as, e.g., Dengue virus, West Nile Virus, Respiratory Syncytial Virus, Epstein-Barr virus, and cytomegalovirus and therefore could play an important role in disease preparedness. Additionally, the VLPNT is a platform technology that could simplify the testing of samples in clinical studies during vaccine development and allow for the immune status surveillance among, e.g., healthcare workers or people at risk.

## Material and Methods

### Cell lines and cell culture

HEK293T (CID3915); HEK293, S^+^ (CID4618); U251MG, hACE2^+^ (CID4663); U251MG, hACE2^+,^ NM∼LgBiT^+^ (CID4697) and Vero, hACE2^+^ (CID4608) cells were maintained in DMEM medium (Gibco, Thermo Fisher Scientific), supplemented with 8% FBS (AC-SM-0143, Anprotec) and Pen/Strep (Gibco, Thermo Fisher Scientific). Cells were cultivated at 37°C in a water-saturated atmosphere with 5% CO_2_. Cell viability was monitored by trypan blue exclusion and cultures with more than 95% viable cells were used in all experiments. Cell lines constitutively expressing S, hACE2 or NM∼LgBiT were generated by retroviral transduction or transient transfection of parental cells with expression plasmids followed by selection with the respective antibiotic and flow cytometric cell sorting.

### SARS-CoV-2 VLPs and other engineered EVs

To generate S^+^ VLPs containing appropriate levels of SARS-CoV-2 spike (D614G or B.1.617.2) and control-EVs, HEK293T cells were transfected using TransIT-293 (MIR2700, Mirus) according to the manufacturers protocol with carefully adjusted ratios of codon-optimized expression vectors coding for S, VSV-G, or mock together with CD63∼HiBiT or CD63∼BlaM and, optionally, M, N and E. The expression plasmids are termed p7413.1 (S:D614G) or p7487.IA1 (S:B.1.617.2), p5451 (VSV-G), p5025 (mock), p7447.SA11 (CD63∼HiBiT), p7200.2 (CD63∼BlaM), p7395.LA3 (M), p7396.NA9 (E), p7391.MA5 (N), respectively.

After 72 hours, S^+^ VLPs, VSV-G^+^ EVs or control EVs were harvested from cell culture supernatants (DMEM, 8% FBS, supplemented with Pen/Strep, Gibco, Thermo Fisher Scientific) after low-speed centrifugation at 7°C for 10 min at 300×g and 20 min at 4200×g and generally used without further processing. If necessary and as indicated, S^+^ VLP and EV supernatants were concentrated by ultrafiltration with 100 kDa cutoff or sedimented by ultra-centrifugation at 100,000×g, 4°C, 2 h followed by floatation in a discontinuous iodixanol (Optiprep, Sigma-Aldrich) density gradient (bottom to top: 1.75 mL sample with 34% final Optiprep concentration; 1.65 mL 30% Optiprep in PBS; 0.6 mL PBS) and ultra-centrifugation at 160,000×g at 4°C for 4 h. Collected fractions were washed with PBS by ultra-centrifugation at 100,000×g, 4°C, 2 h, and resuspended. Particle preparations were flash frozen in liquid nitrogen for storage at −80°C and subsequently characterized by WB, NTA, nano flow technology, ELISA and cryo-EM.

### Western blot

Concentrated S^+^ VLPs or EVs were lysed in non-reducing 5× Laemmli buffer and separated by SDS-PAGE. Proteins were transferred to nitrocellulose membranes (GE Healthcare Life Science) by semi-dry blotting (Bio-Rad) and membranes were blocked for 1 h in 5% (w/v) non-fat milk in ddH_2_O at room temperature (RT). Membranes were incubated at 7°C overnight with primary anti-spike antibody (43A11, rat IgG, Helmholtz Zentrum Mu□nchen; 1A9, mouse IgG, GeneTex; PA5-81795, polyclonal rabbit antibody, Invitrogen) at 1:2000 dilution in 5% (w/v) non-fat milk (Carl Roth) in ddH_2_O, washed 3 times in TBST (Tris-buffered saline with 0.1% Tween-20) and incubated at RT for 1 h with horseradish peroxidase (HRP)-conjugated secondary antibody (1112-035-062, goat anti-rat IgG, Jackson Immuno Research Europe; 7076S, horse anti-mouse IgG, Cell Signaling; 7074S, goat anti rabbit IgG, Cell Signaling) at 1:20,000, 1:2000 and 1:2000 dilution respectively, in 5% (w/v) non-fat milk in PBST (PBS, 0.05% Tween-20). After repeated (3×) washing in TBST, blots were incubated with ECL reagent (GE Healthcare) and imaged using a Fusion FX (Vilber).

### ELISA

For the quantitation of spike in samples from various sources, a sandwich enzyme-linked immunosorbent assay (ELISA) was developed, using two anti-spike antibodies with non-overlapping epitopes. First, wells of a Nunc MaxiSorp plate (Thermo Fisher Scientific) were coated at RT for 5 h with 2 µg mL^-1^ anti-spike capture antibody (55E10, rat IgG, Helmholtz Zentrum Mu□nchen) or isotype in PBS. After washing with PBST (PBS, 0.05% Tween-20), free binding sites were blocked at RT for 2 h in 5% (w/v) non-fat milk (Carl Roth) in PBS. Samples of recombinant spike protein (S1+S2 extracellular domain, 40589-V08B1, Sino Biological), S^+^ VLPs, or controls were diluted as indicated (Fig. 3A, B) in PBS and incubated at 7°C for 16 h on the antibody coated plate. After washing, HRP conjugated anti-spike detection antibody (43A11, rat IgG, Helmholtz Zentrum Mu□nchen) diluted 1:500 in 5% (w/v) non-fat milk in PBS) was added at RT for 2 h, washed again and incubated at RT with 100 µL TMB substrate reagent (BD555214, Becton Dickinson). The reaction was stopped by adding 50 µL 1 M H_2_SO_4_ and absorbance was measured at 450 nm in a CLARIOstar Plus (BMG Labtech). Data analysis was performed with GraphPad Prism 9.2.

### NTA Analysis

Nanoparticle tracking analysis (NTA) was performed with the ZetaView PMX110 instrument (Particle Metrix) and the corresponding software (ZetaView 8.04.02) was used to measure the number and the size distribution of the S^+^ VLP, virus stock and EV preparations. Samples were diluted in filtered PBS to achieve a vesicle concentration of approximately 1×10^7^ mL^-1^. Pre-acquisition parameters were set to a sensitivity of 75, a shutter speed of 50, a frame rate of 30 frames per second and a trace length of 15. The post-acquisition parameters were set to a minimum brightness of 20, a minimum size of 5 pixels and a maximum size of 1000 pixels.

### Analysis of S^+^ VLP, SARS-CoV-2 and EV particles with nano flow technology

Single particles were analyzed using our nano flow technology and a CytoFLEX LX cytometer (Beckman Coulter Life Science) with samples of S^+^ VLPs, SARS-CoV-2 stock and EVs pre-stained with CellTraceViolet (CTV, Thermo Fisher Scientific) at 37°C for 20 min and quenched with PBS with 1% BSA. Upon washing via a 100 kDa cutoff Amicon ultra filter, the samples were stained with AlexaFluor488 conjugated anti-S antibody (43A11, rat IgG, Helmholtz Zentrum Mu□nchen), diluted and afterwards analyzed in the flow cytometer.

### Cryo-electron microscopy

Aliquots of 4 μL from crude or purified S^+^ VLPs were deposited on EM grids coated with a perforated carbon film. After draining the excess liquid with a filter paper, grids were quickly plunged into liquid ethane cooled by liquid nitrogen, using a Leica EMCPC cryo-chamber, and stored in cryo-boxes under liquid nitrogen until use. For cryo-EM observation, grids were mounted onto a Gatan 626 cryoholder and transferred to a Tecnai F20 microscope (FEI, USA) operated at 200 kV. Images were recorded with an Eagle 2k CCD camera (Thermo Fisher, USA).

### EV fusion assay with BlaM readout

For EV fusion or EV neutralization assays with cytometric readouts, 2×10^4^ Vero, hACE2^+^ or U251MG, hACE2^+^ cells (CID4608, CID4663, Helmholtz Zentrum München) in 100 µL DMEM (8% FBS, Pen/Strep supplemented) were seeded per well in a 96-well plate (353072, Falcon) and incubated o/n at 37°C, 5% CO_2_. Where indicated, the cells were pre-incubated at 37°C for 1 h with chloroquine or were left without further treatment. The spent medium was replaced with up to 100 µL concentrated EVs (S^+^, mock or VSV-G^+^, CD63∼BlaM^+^) with or without prior pre-incubation with serum samples. After 4 h incubation at 37°C, 5% CO_2_, the supernatant was removed, adherent cells were washed in PBS, trypsinized and moved to a V-bottom shaped 96-well plate and sedimented by centrifugation. After washing the cells once in CO_2_ independent medium (8% FBS, Pen/Strep supplemented, Gibco, Thermo Fisher Scientific), 50 µL staining solution were added and the cells were incubated at RT in the dark o/n. The staining solution was composed of 2 µL CCF4-AM, 8 µL solution B (K1095, Thermo Fisher Scientific) and 10 µL 250 mM Probenecid (P8761, Sigma) per 1 mL supplemented CO_2_ independent medium. After enzymatic turnover of the CCF4, cells were washed once with PBS (+3% FBS) and analyzed by flow cytometry using an LSR Fortessa instrument (BD) with a high throughput auto sampler. The 409-nm wavelength laser (violet) was used for excitation of the FRET substrate, resulting in emission of intact, non-cleaved CCF4 substrate at 520 nm (green), whereas emission of cleaved CCF4 substrate was detected at 447 nm (blue).

### VLPNT with nanoLuciferase readout

For virus-like-particle neuralization tests (VLPNTs), 2×10^4^ U251MG (hACE2^+^, NM∼LgBiT^+^) cells (CID4697, Helmholtz Zentrum München) in 100 µL DMEM (8% FBS, Pen/Strep supplemented) were seeded per well in a 96-well Lumitrac200 (655075, Greiner Bio-One) plate and incubated at 37°C, 5% CO_2_ overnight. The following day, serial dilutions of serum samples by a factor of 2 starting at 1:2 to 1:360 were prepared in PBS in a 96-well plate. 12 µL of the serial serum dilutions each were added to 48 µL S^+^ VLP (S, M, N, E, CD63∼HiBiT) to obtain a series of final dilutions ranging from 1:10 to 1:1800. S^+^ VLPs and serum dilutions were incubated at 37°C for 30 min in a 96-well plate. Thereafter, the mix was transferred to recipient cells after removal of their culture medium and the cells were incubated at 37°C, 5% CO_2_ for 4 h. Prior to readout, the supernatant was removed and replaced with 25 µL substrate mix (20 µL OptiMEM, Gibco, Thermo Fisher Scientific +5 µL nano-Glo diluted 1:20 in LCS buffer, N2012, Promega). Bioluminescence was immediately quantified in a CLARIOstar Plus reader (BMG Labtech). Mean luminescence level of S^+^ VLPs, only, was set to 0% neutralization; while background luminescence obtained with Δ vFP EVs was set to 100% neutralization. The half maximal neutralization titer was determined by extrapolating sigmoidal curve values that correspond to 50% signal reduction after background correction. This value was termed VLPN_50_. The WHO international standard (NIBSC 20/136) was used in between runs and as within-run reference.

### Anti-spike antibody generation

To obtain spike antigen in its authentic conformation for the immunization of rats, a constitutive spike expressing HEK293 cell line (CID4618, Helmholtz Zentrum Mu□nchen) expressing SARS-CoV-2 spike^FL^ (D614G) was established. To boost expression levels further, the cells were transiently transfected with an S expression plasmid (p7413, Helmholtz Zentrum Mu□nchen) prior to the isolation of S^+^ EVs from conditioned media three days after transfection. EVs were purified and concentrated by ultrafiltration, density gradient ultra-centrifugation and subsequently characterized as described above. For the generation of anti-spike antibodies, S^+^ EVs were used to immunize rats in a prime-boost-boost scheme, before splenic B-cells were isolated to establish hybridoma cell clones. Specific clones were subsequently identified by screening of hybridoma supernatants for binding to the constitutive spike expressing HEK293 cell line (CID4618, Helmholtz Zentrum Mu□nchen) via flow cytometry. The resulting anti-spike antibodies (COVEV: 43A11 and 55E10) were then characterized thoroughly to validate their high affinity and specificity for spike^FL^.

### Antibody production

Antibodies 43A11 and 55E10 were purified from hybridoma supernatants (Helmholtz Zentrum Mu□nchen Core Facility Monoclonal Antibodies). Antibodies REGN10987 and REGN10933 were purified from CHO cells transiently transfected with expression plasmids encoding the heavy and light chains. Briefly, cell culture supernatant was loaded on a protein GammaBind Plus Sepharose (17088602, Cytiva) or HiTrap MabSelect SuRe (29049104, Cytiva) column, washed with PBS and antibodies were eluted at pH 2.7 or 3.5, respectively in citric acid buffer. Fractions were pooled, neutralized with 1 M Tris (pH 8.0) and separated on Superdex200 prep grade (17104301, Cytiva) in PBS for monomers or buffer exchanged to PBS via ultra-filtration, respectively. Directly coupled AlexaFluor488 labelled antibodies were generated using commercial kits (A10235, Thermo Fisher Scientific).

### SARS-CoV-2 pseudotyped virus neutralization test (pVNT)

S-pseudotyped retrovirus stocks were generated by co-transfecting expression plasmids coding for S, gag and pol (from murine leukemia virus) and a myeloproliferative sarcoma virus (MPSV) based retroviral vector encoding eGFP (p7413, p4037, p6895, Helmholtz Zentrum Mu□nchen) into HEK293T cells. After 48 h, the supernatant was harvested and centrifuged at 7°C for 10 min at 300×g, 20 min at 4200×g and flash frozen for storage (−80°C). For pVNTs, 5×10^3^ Vero (hACE2^+^) cells (CID4608, Helmholtz Zentrum München) in 100 µL DMEM (8% FBS, Pen/Strep supplemented) were seeded per well in a 96-well plate (353072, Falcon) and incubated at 37°C, 5% CO_2_ overnight. The cells were then pre-incubated for 1 h at 37°C in 50 µL, 8 µg mL^-1^ protamine sulfate in DMEM with or without chloroquine and retroviral stocks were pre-incubated for 30 min at 37°C with 5× serum dilutions in PBS. 50 µL of this mix where then added to the pre-incubated cells and centrifuged for 90 min at 800×g, at 37°C. After 48 h at 37°C, GFP expression was quantified by flow cytometry using an LSR Fortessa instrument (BD).

### SARS-CoV-2 neutralization test (VNT_100_)

Human sera were heat-inactivated at 56°C for 30 min and diluted in a series of two-fold dilution steps (1:4 to 1:512). 100 plaque-forming units (PFU) of SARS-CoV-2 stock (German isolate BavPat1/2020; European Virus Archive Global # 026 V-03883, GenBank: MZ558051.1) contained in 50µL was added to an equal volume of diluted serum. The mixture was incubated at 37°C and approximately 20,000 Vero C1008 cells (ATCC, Cat#CRL-1586) were added after 1 hour. After a four-day incubation period, the cytopathic effect (CPE) was evaluated by light microscopy. Virus neutralization was defined as the complete absence of CPE in a given serum dilution (VNT_100_). The reciprocal geometric mean titer (GMT) was calculated from the highest serum dilution without CPE based on three replicates. The lower detection limit of the assay was 1:8, corresponding to the first dilution of the serum tested. Two positive controls were used as inter-assay neutralization standards and quality control for each test. The WHO international standard (NIBSC 20/136) was tested seven times, resulting in a GMT of 377 for 100% absence of CPE in this test. Neutralization assays were performed in the BSL-4 laboratory of the Institute of Virology at Philipps University Marburg, Germany (Romero-Olmedo et al., 2021).

### Patients and specimens

We included 23 serum specimens collected between April 6, 2020, and June 25, 2020, from 18 patients infected with SARS-CoV-2 at the LMU Klinikum, Munich, Germany. Patients are part of the COVID-19 Registry of the LMU Klinikum (CORKUM, WHO trial id DRKS00021225) and the study was approved on March 23, 2020 by the ethics committee (no. 20-245) of the Faculty of Medicine of the LMU (Ethik-Kommission bei der Medizinischen Fakultät der Ludwig-Maximilians-Universität München, Pettenkoferstr. 8a, 80336 München, Germany). Clinical data were obtained from health records and all patient data were anonymized for analysis. All patients were tested positive for SARS-CoV-2 by rRT-PCR in nasopharyngeal or oropharyngeal swabs. The median age of the 18 COVID-19 patients was 60 years (interquartile range 52 to 75 years), and 11.1% (2/18) of these individuals were female. We categorized the disease severity of the COVID-19 patients according to the WHO guideline “Clinical Management of COVID-19” (World Health Organization, 2020): asymptomatic (no clinical signs of infection), mild (symptomatic patients without evidence of viral pneumonia or hypoxia), moderate (clinical signs of pneumonia, including fever, cough, dyspnoea), severe (clinical signs of pneumonia, plus one of the following: respiratory rate >30 min^-1^, severe respiratory distress, SpO_2_ <90% on room air), critical (one of the following: acute respiratory distress syndrome, sepsis, septic shock). Three patients were categorized as asymptomatic, 2 patients as mild, 6 patients as moderate, 5 patients as severe, and 2 patients as critical.

### Virus stock preparation

CaCo-2 cells (American Type Culture Collection, ATCC, Virginia, USA) in cell culture medium (Dulbecco’s Modified Eagle’s Medium containing 2% FBS) were challenged for 2 h with a clinical isolate of SARS-CoV-2 (GISAID EPI ISL 2967222) previously obtained from a nasopharyngeal swab of a COVID-19 patient. Subsequently, cell culture medium was exchanged, and three days post infection supernatants were passaged on Vero-E6 cells (ATCC, Virginia, USA). After three additional days, the cell culture supernatant was harvested and stored at −80°C. The virus stock was characterized by rRT-PCR. Heat-inactivation of the virus stock was performed by incubating the sample for 30 min at 56°C.

## Data Availability

All data produced in the present work are contained in the manuscript

## Acknowledgements

We would like to thank Regina Feederle and her team, Helmholtz Zentrum München, Core Unit Monoclonal Antibodies and Markus Kellner, Helmholtz Zentrum München, Therapeutic Antibody Unit, for providing antibodies crucial for this work. We would like to thank Marcel Stern, Max von Pettenkofer Institute, LMU München for establishing the SARS-CoV-2 virus stock used. We would like to thank Sisareuth Tan, UMR-CBMN CNRS-University of Bordeaux-INP, for his expert technical assistance in cryo-EM and Percy Knolle and Bastian Höchst, Technical University of Munich for arranging the collaboration with Alain Brisson. We would like to thank all CORKUM investigators and staff. The authors thank the patients and their families for their participation in the CORKUM registry. We acknowledge the European Virus Archive Global (EVAg) for providing the virus isolate used in this study for the cVNT. We would like to thank Ronan Le Gleut, Helmholtz Zentrum München, Core Facility Statistical Consulting, for consultation on statistics. This research project was supported by LMUexcellent, funded by the Federal Ministry of Education and Research (BMBF) and the Free State of Bavaria under the Excellence Strategy of the Federal Government and the Länder. This work was also financially supported by grants from the Deutsche Forschungsgemeinschaft (grant numbers SFB1064/TP A13, SFB-TR36/TP A04) and Deutsche Krebshilfe (grant number 70112875) to W.H. and Bundesministerium für Bildung und Forschung (grant number BMBF 01KI2040) to R.Z..

## Author contributions

JR performed most of the experiments; DP supported by WH provided substantial additional experimental work; MA and OK provided the SARS-CoV-2 virus stock; PR, OK, JH, CS, and MB provided the 23 clinical COVID-19 samples; VK and SB performed cVNTs; AB performed cryo-EM; JR and WH wrote the paper; JR, RZ and WH designed the scientific concept and the experimental realization.

## Competing interests

The authors declare no competing interests.

## Supporting information

**Supplementary Figure S1.**
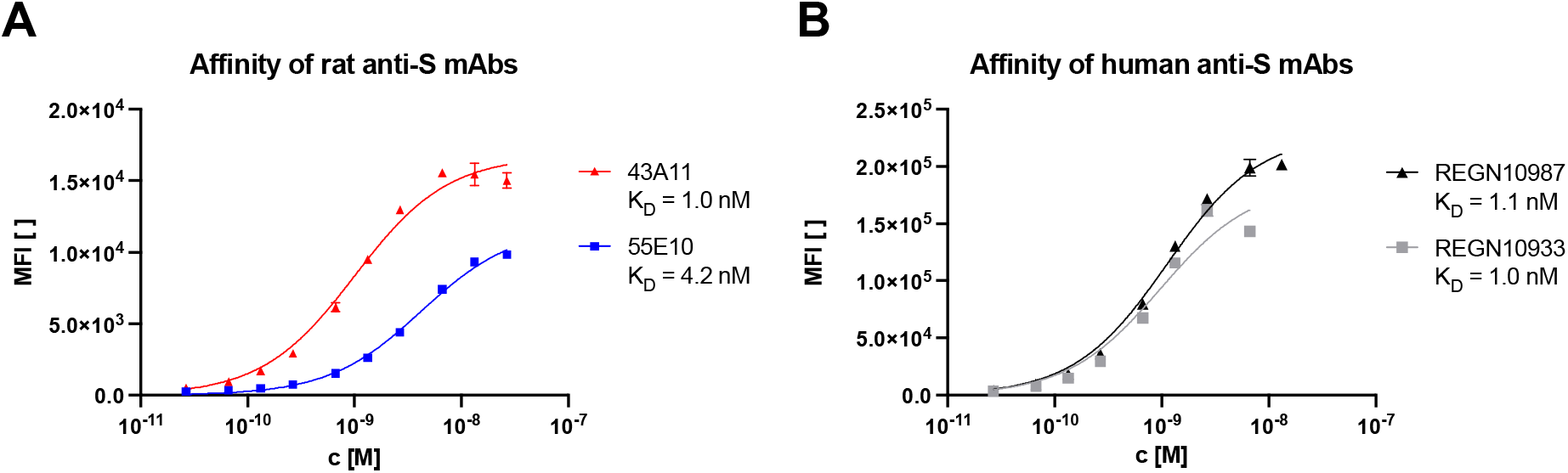
Binding and dissociation constant estimation of four spike-specific mAbs. Binding affinities of four recombinant anti-spike mAbs were analyzed by flow cytometry using a HEK293 cell line that expresses spike protein constitutively. Mean fluorescence intensity (MFI) values were determined for various concentrations (c) of the antibodies with the aid of a labeled secondary antibody. MFI data of biological triplicates were analyzed according to a one-site specific binding model to estimate the dissociation constant K_D_ of the mAb to its antigen. (A) Binding curves of two in house generated rat IgG anti-S mAbs 43A11 and 55E10 are shown. (B) Binding curves of two spike-specific neutralizing human IgG mAbs, REGN10987 and REGN10933, are shown for comparison.

**Supplementary Figure S2.**
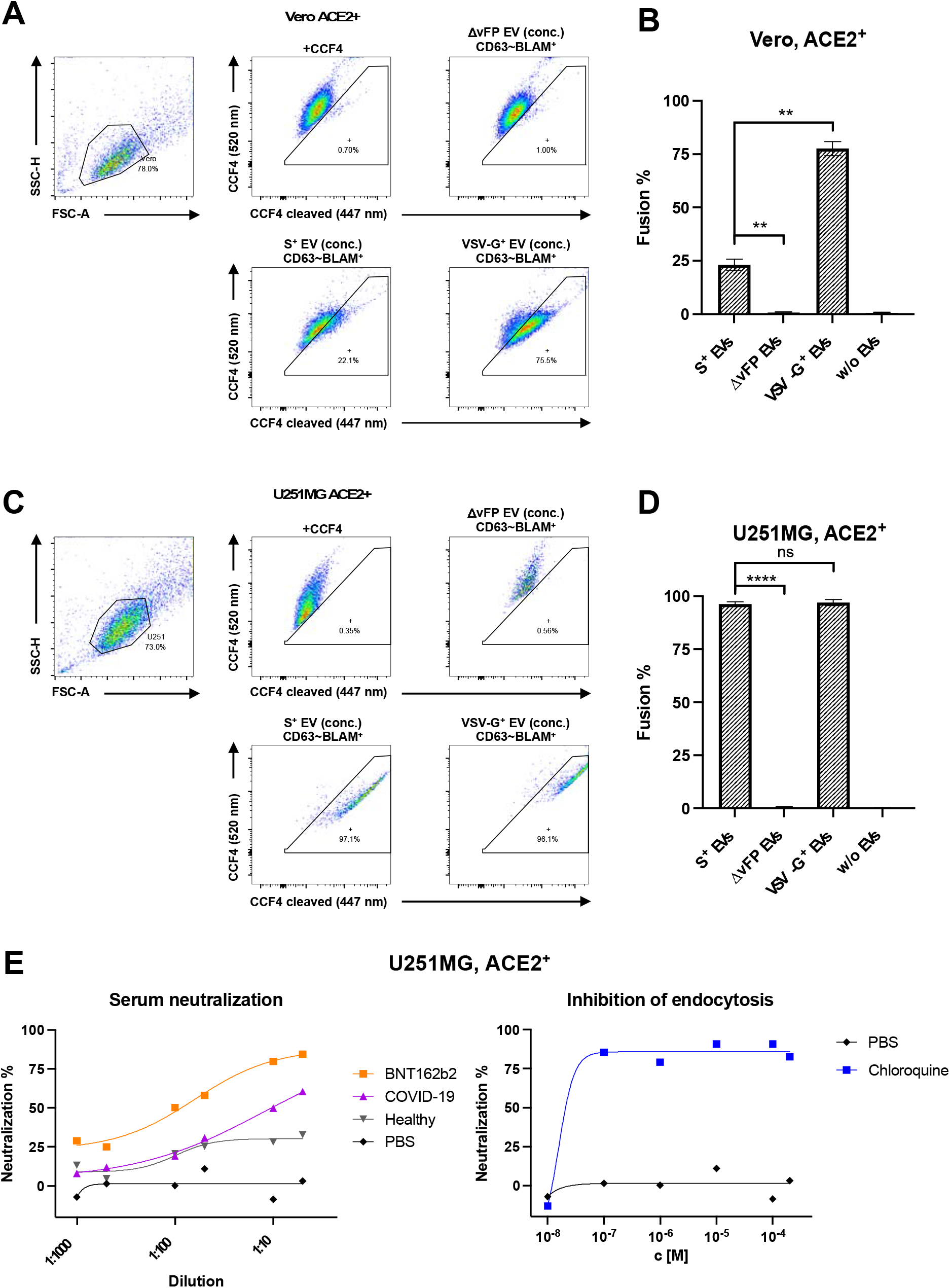
VLPNT with the BlaM reporter system. A version of the spike-specific VLPNT is shown which uses extracellular vesicles (EVs) and β-lactamase (BlaM) as protein reporter. It consists of a fusion of BlaM to the carboxy-terminus of human CD63, which is efficiently incorporated into the membrane of EVs. HEK293T cells transiently co-transfected with CD63∼BlaM and spike encoding plasmids release extracellular vesicles termed S^+^ EVs. Upon incubation of ACE2^+^ recipient cells, CD63∼BlaM is transferred to the cells’ cytoplasm and BlaM cleaves the substrate CCF4 altering the emission spectrum of the cells, which is analyzed by flow cytometry. BlaM-positive cells within the gate show blueish fluorescence (x-axis) compared with greenish cells, which contain uncleaved CCF4 (y-axis). (A) Flow cytometry plots show the gating strategy of intact Vero cells (left panel) as recipient cells, engineered to express human ACE2 and loaded with the BlaM substrate CCF4. The top row middle and right panels show untreated Vero hACE2^+^ cells and cells incubated with concentrated (conc.) Δ vFP EVs, respectively. The Δ vFP EVs contain CD63∼BlaM, only, but lack the viral fusion protein (vFP). The two bottom panels show examples of recipient Vero hACE2^+^ cells after the successful delivery of ß-lactamase using two different types of particles. The lower left and right panels show Vero ACE2^+^ cells incubated with S^+^ EVs and VSV-G^+^ EVs, respectively. The fraction of cells which have taken up VSV-G^+^ EVs is larger compared to the fraction of cells which were incubated with S^+^ EVs. (B) Data from four biological replicates (examples are shown in panel A) are summarized and graphically displayed. Results from t-tests are indicated; ns, not significant (p > 0.05); *p ≤ 0.05; **p ≤ 0.01; ***p ≤ 0.001; ****p ≤ 0.0001. (C, D) Experiments were conducted as in panels A and B with U251MG recipient cells stably expressing human ACE2. They were found to be better targets for S^+^ EVs than ACE2^+^ Vero cells as 97% of the ACE2^+^ U251MG cells incubated with S^+^ EV contained cleaved CCF4 compared to 21.4% of ACE2^+^ Vero cells in panels A and B. (E) Neutralization experiments with three sera and a PBS control using ACE2^+^ U251MG cells as recipients and S^+^ EVs containing the CD63∼BlaM protein reporter. In the left panel, concentration dependent neutralization of S^+^ EVs using sera from an individual vaccinated with BNT162b2 and a COVID-19 convalescent are compared with a naïve, healthy donor (and PBS as negative control). Fusion of S^+^ EVs with ACE2^+^ U251MG cells was efficiently blocked by low concentrations (c) of chloroquine indicative of the endosomal uptake of S^+^ EVs (right panel).

**Supplementary Figure S3.**
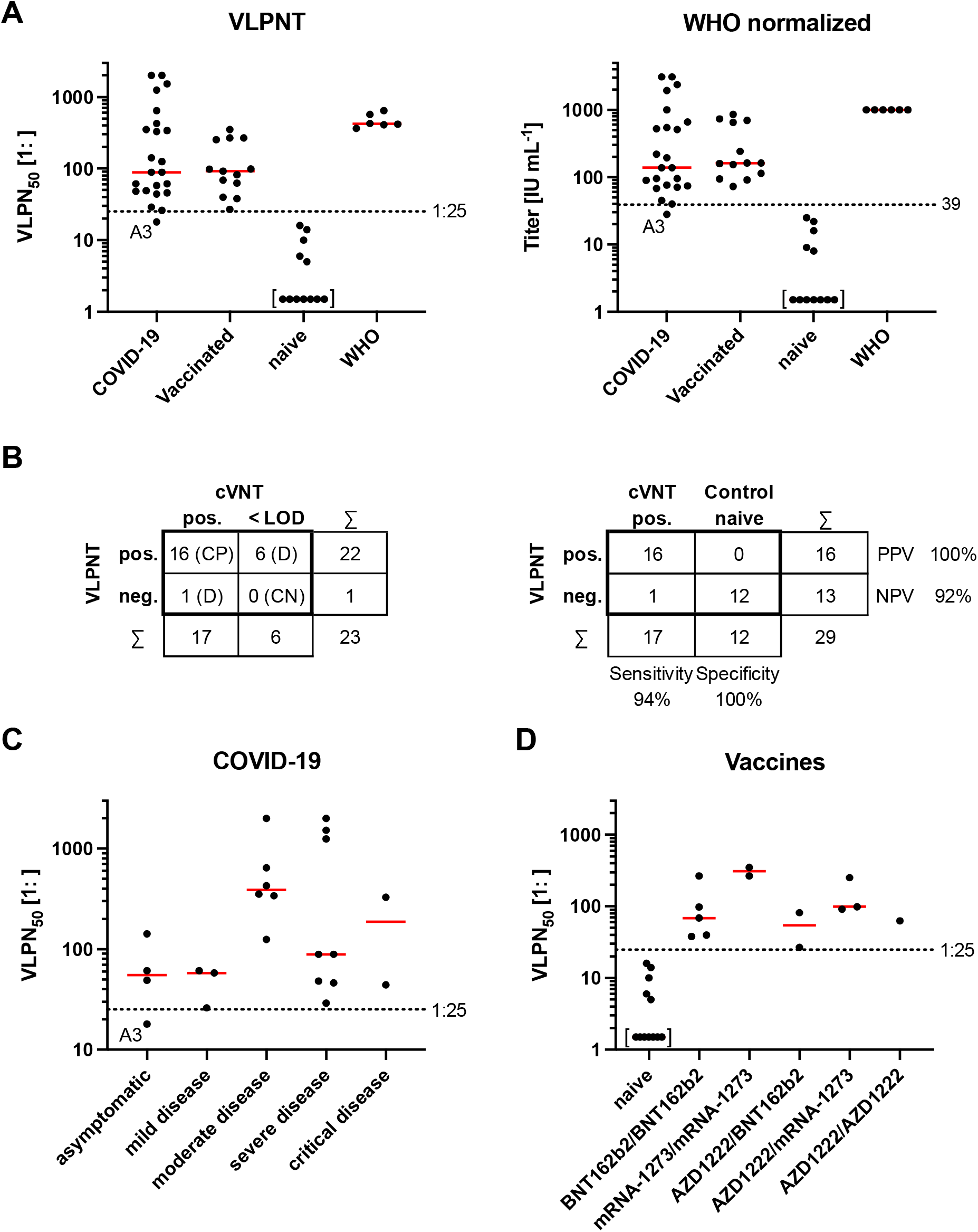
Diagnostic VLPNT performance and serum titer classification according to the donors’ medical history. (A) VLPN_50_ titers of sera from 23 COVID-19 patients, 13 COVID-19 vaccinees, 12 healthy naïve donors and six independent replicates of the WHO convalescent COVID-19 reference serum (NIBSC 20/136) are grouped and shown with median values (red bars). The dashed horizontal line indicates VLPN_50_ titers below which the test does not score spike-specific neutralizing antibodies according to our criteria. The left and right panels show the grouped VLPN_50_ titers prior to and after their normalization to international units (IU) with the WHO standard, respectively. Values in square brackets indicate sera with VLPN_50_ values below the limit of detection (LOD). The sample denoted A3 is a single serum sample referred to in the Result section. (B) The left panel compares the results of 23 COVID-19 samples tested in the VLPNT and the cVNT. Concordant positive (CP), concordant negative (CN), and discrepant (D) results are indicated. In the right panel, sensitivity, specificity, positive predictive value (PPV) and negative predictive value (NPV) of the VLPNT were calculated based on the 17 COVID-19 serum samples with neutralizing capacity in the cVNT and 12 serum controls of healthy naïve donors collected prior to mid 2019. Test results are indicated as: pos., positive; neg., negative; < LOD, below LOD. (C) VLPN_50_ serum titers of 23 COVID-19 patients are sorted according to their clinical disease course. Red bars indicate median values of grouped data. (D) VLPN_50_ serum titers of vaccinees are shown according to their prime-boost scheme of different COVID-19 vaccines received. Red bars indicate median values of the groups. Values in square brackets indicate serum samples with VLPN_50_ below the LOD.

**Supplementary Figure S4.**
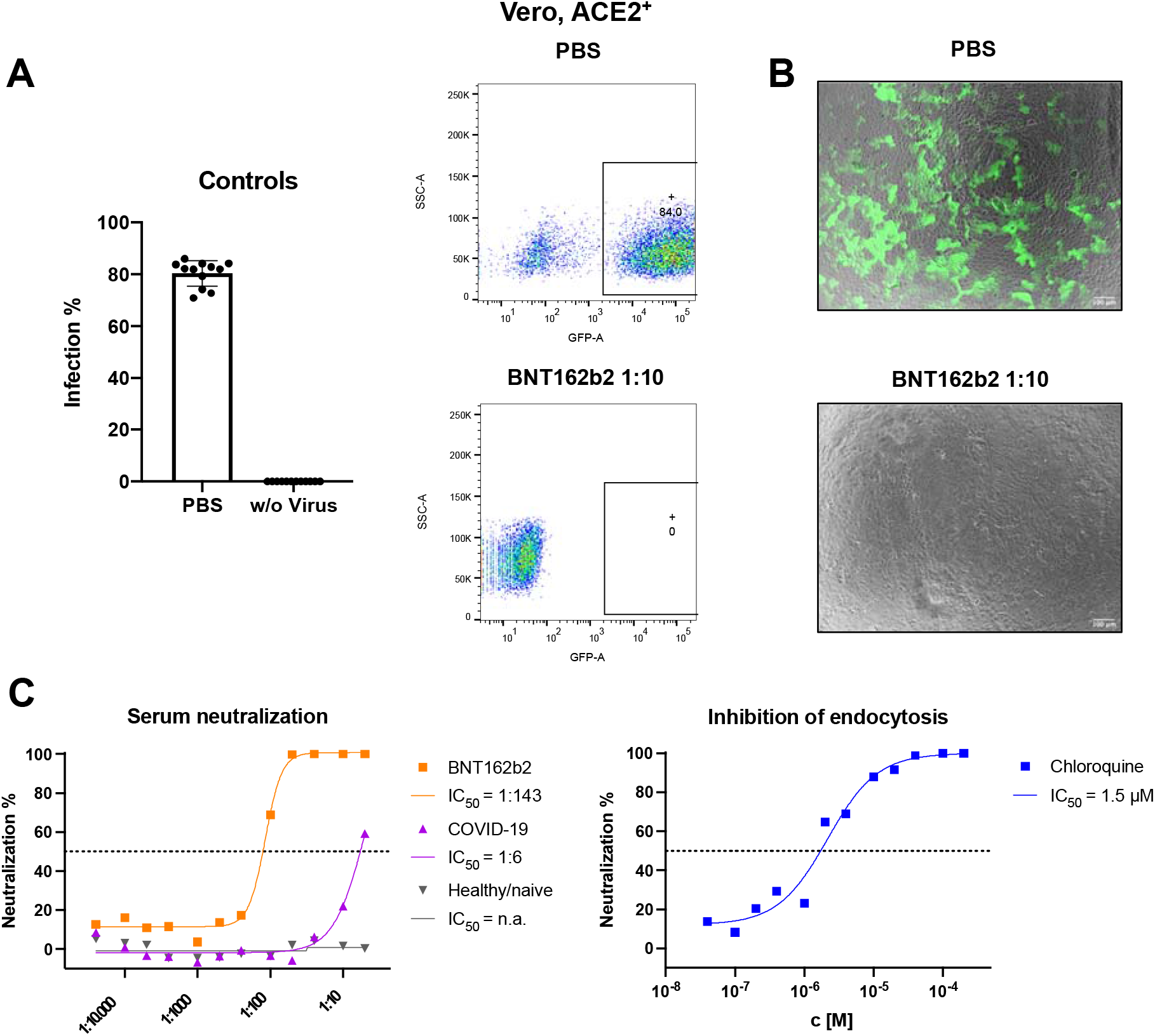
pVNT with spike-pseudotyped retroviral vectors encoding GFP. Spike-pseudotyped retrovirus stock based on a murine leukemia virus vector encoding GFP was used in pseudotyped virus neutralization tests (pVNT) for the quantification of NAbs. Readouts rely on *de novo* expression of the retrovirally transduced GFP gene in ACE2^+^ recipient cells analyzed by flow cytometry. (A) A spike-pseudotyped retroviral vector preparation was incubated with Vero cells engineered to express human ACE2. The cells of 12 biological replicates were analyzed for GFP expression 48 h post infection by flow-cytometry yielding about 80% GFP^+^ cells (top dot plot). Preincubation of the retroviral vector inoculum with a final concentration of a 1:10 diluted serum from a BNT162b2 vaccinated individual for 30 min efficiently blocked GFP expression (bottom plot). (B) Merged fluorescence and brightfield microscopic images of Vero ACE2^+^ cells visualize recipient cells infected with spike-pseudotyped retroviral vectors incubated with PBS as a negative control (top) and after adding neutralizing serum (bottom) as examples of the experiment described in panel A. White scale bar corresponds to 100 µm in length. (C) Three sera from a vaccinee after BNT162b2 prime-boost immunization, a COVID-19 convalescent and a healthy naïve donor were titrated with fixed doses of spike-pseudotyped retroviral vector inoculum (left panel). Infection of ACE2^+^ Vero cells with pseudotyped vector particles was efficiently blocked by pre-incubating recipient cells with low concentrations (c) of chloroquine preventing endocytosis (right panel). Half maximal inhibitory concentrations IC_50_ are calculated, or indicated as not applicable (n.a.).

## Notes

### Competing Interest Statement

The authors have declared no competing interest.

### Author Declarations

Patients are part of the COVID-19 Registry of the LMU Klinikum (CORKUM, WHO trial id DRKS00021225) and the study was approved on March 23, 2020 by the Ethics committee (no. 20-245) of the Faculty of Medicine of the LMU (Ethik-Kommission bei der Medizinischen Fakultaet der Ludwig-Maximilians-Universitaet Muenchen, Pettenkoferstr. 8a, 80336 Muenchen, Germany)

## References

Anonymous (2020). An EUA for casirivimab and imdevimab for COVID-19. Med Lett Drugs Ther 62, 201–202.

Abu-Raddad, L.J., Chemaitelly, H., Butt, A.A., and National Study Group for, C.-V. (2021). Effectiveness of the BNT162b2 Covid-19 Vaccine against the B.1.1.7 and B.1.351 Variants. N Engl J Med 385, 187–189. DOI:10.1056/NEJMc2104974

Albanese, M., Chen, Y.A., Huls, C., Gartner, K., Tagawa, T., Mejias-Perez, E., Keppler, O.T., Gobel, C., Zeidler, R., Shein, M., et al. (2021). MicroRNAs are minor constituents of extracellular vesicles that are rarely delivered to target cells. PLoS Genet 17, e1009951. DOI:10.1371/journal.pgen.1009951

Baum, A., Fulton, B.O., Wloga, E., Copin, R., Pascal, K.E., Russo, V., Giordano, S., Lanza, K., Negron, N., Ni, M., et al. (2020). Antibody cocktail to SARS-CoV-2 spike protein prevents rapid mutational escape seen with individual antibodies. Science 369, 1014–1018. DOI:10.1126/science.abd0831

Brouwer, P.J.M., Caniels, T.G., van der Straten, K., Snitselaar, J.L., Aldon, Y., Bangaru, S., Torres, J.L., Okba, N.M.A., Claireaux, M., Kerster, G., et al. (2020). Potent neutralizing antibodies from COVID-19 patients define multiple targets of vulnerability. Science 369, 643–650. DOI:10.1126/science.abc5902

Burki, T.K. (2021). Lifting of COVID-19 restrictions in the UK and the Delta variant. Lancet Respir Med 9, e85. DOI:10.1016/S2213-2600(21)00328-3

Carlon-Andres, I., and Padilla-Parra, S. (2020). Quantitative FRET-FLIM-BlaM to Assess the Extent of HIV-1 Fusion in Live Cells. Viruses 12. DOI:10.3390/v12020206

Cavrois, M., De Noronha, C., and Greene, W.C. (2002). A sensitive and specific enzyme-based assay detecting HIV-1 virion fusion in primary T lymphocytes. Nat Biotechnol 20, 1151–1154. DOI:10.1038/nbt745

Cavrois, M., Neidleman, J., and Greene, W.C. (2014). HIV-1 Fusion Assay. Bio Protoc 4. DOI:10.21769/bioprotoc.1212

Chesher, D. (2008). Evaluating assay precision. Clin Biochem Rev 29 Suppl 1, S23–26.

Cohen, K.W., Linderman, S.L., Moodie, Z., Czartoski, J., Lai, L., Mantus, G., Norwood, C., Nyhoff, L.E., Edara, V.V., Floyd, K., et al. (2021). Longitudinal analysis shows durable and broad immune memory after SARS-CoV-2 infection with persisting antibody responses and memory B and T cells. Cell Rep Med 2, 100354. DOI:10.1016/j.xcrm.2021.100354

Davies, N.G., Abbott, S., Barnard, R.C., Jarvis, C.I., Kucharski, A.J., Munday, J.D., Pearson, C.A.B., Russell, T.W., Tully, D.C., Washburne, A.D., et al. (2021). Estimated transmissibility and impact of SARS-CoV-2 lineage B.1.1.7 in England. Science 372. DOI:10.1126/science.abg3055

DeFrancesco, L. (2020). COVID-19 antibodies on trial. Nat Biotechnol 38, 1242–1252. DOI:10.1038/s41587-020-0732-8

Desai, T.M., Marin, M., Mason, C., and Melikyan, G.B. (2017). pH regulation in early endosomes and interferon-inducible transmembrane proteins control avian retrovirus fusion. J Biol Chem 292, 7817–7827. DOI:10.1074/jbc.M117.783878

Dixon, A.S., Schwinn, M.K., Hall, M.P., Zimmerman, K., Otto, P., Lubben, T.H., Butler, B.L., Binkowski, B.F., Machleidt, T., Kirkland, T.A., et al. (2016). NanoLuc Complementation Reporter Optimized for Accurate Measurement of Protein Interactions in Cells. ACS Chem Biol 11, 400–408. DOI:10.1021/acschembio.5b00753

Earle, K.A., Ambrosino, D.M., Fiore-Gartland, A., Goldblatt, D., Gilbert, P.B., Siber, G.R., Dull, P., and Plotkin, S.A. (2021). Evidence for antibody as a protective correlate for COVID-19 vaccines. Vaccine 39, 4423–4428. DOI:10.1016/j.vaccine.2021.05.063

Feeley, E.M., Sims, J.S., John, S.P., Chin, C.R., Pertel, T., Chen, L.M., Gaiha, G.D., Ryan, B.J., Donis, R.O., Elledge, S.J., et al. (2011). IFITM3 inhibits influenza A virus infection by preventing cytosolic entry. PLoS Pathog 7, e1002337. DOI:10.1371/journal.ppat.1002337

Feng, S., Phillips, D.J., White, T., Sayal, H., Aley, P.K., Bibi, S., Dold, C., Fuskova, M., Gilbert, S.C., Hirsch, I., et al. (2021). Correlates of protection against symptomatic and asymptomatic SARS-CoV-2 infection. Nat Med 27, 2032–2040. DOI:10.1038/s41591-021-01540-1

Giroglou, T., Cinatl, J., Jr., Rabenau, H., Drosten, C., Schwalbe, H., Doerr, H.W., and von Laer, D. (2004). Retroviral vectors pseudotyped with severe acute respiratory syndrome coronavirus S protein. J Virol 78, 9007–9015. DOI:10.1128/JVI.78.17.9007-9015.2004

Hoffmann, M., Kleine-Weber, H., Schroeder, S., Kruger, N., Herrler, T., Erichsen, S., Schiergens, T.S., Herrler, G., Wu, N.H., Nitsche, A., et al. (2020a). SARS-CoV-2 Cell Entry Depends on ACE2 and TMPRSS2 and Is Blocked by a Clinically Proven Protease Inhibitor. Cell 181, 271–280 e278. DOI:10.1016/j.cell.2020.02.052

Hoffmann, M., Mosbauer, K., Hofmann-Winkler, H., Kaul, A., Kleine-Weber, H., Kruger, N., Gassen, N.C., Muller, M.A., Drosten, C., and Pohlmann, S. (2020b). Chloroquine does not inhibit infection of human lung cells with SARS-CoV-2. Nature 585, 588–590. DOI:10.1038/s41586-020-2575-3

Hofmann, H., and Pohlmann, S. (2004). Cellular entry of the SARS coronavirus. Trends Microbiol 12, 466–472. DOI:10.1016/j.tim.2004.08.008

Hsieh, P.K., Chang, S.C., Huang, C.C., Lee, T.T., Hsiao, C.W., Kou, Y.H., Chen, I.Y., Chang, C.K., Huang, T.H., and Chang, M.F. (2005). Assembly of severe acute respiratory syndrome coronavirus RNA packaging signal into virus-like particles is nucleocapsid dependent. J Virol 79, 13848–13855. DOI:10.1128/JVI.79.22.13848-13855.2005

Hu, B., Guo, H., Zhou, P., and Shi, Z.L. (2021). Characteristics of SARS-CoV-2 and COVID-19. Nat Rev Microbiol 19, 141–154. DOI:10.1038/s41579-020-00459-7

Jones, D.M., and Padilla-Parra, S. (2016). The beta-Lactamase Assay: Harnessing a FRET Biosensor to Analyse Viral Fusion Mechanisms. Sensors (Basel) 16. DOI:10.3390/s16070950

Ke, Z., Oton, J., Qu, K., Cortese, M., Zila, V., McKeane, L., Nakane, T., Zivanov, J., Neufeldt, C.J., Cerikan, B., et al. (2020). Structures and distributions of SARS-CoV-2 spike proteins on intact virions. Nature 588, 498–502. DOI:10.1038/s41586-020-2665-2

Keng, C.T., Zhang, A., Shen, S., Lip, K.M., Fielding, B.C., Tan, T.H., Chou, C.F., Loh, C.B., Wang, S., Fu, J., et al. (2005). Amino acids 1055 to 1192 in the S2 region of severe acute respiratory syndrome coronavirus S protein induce neutralizing antibodies: implications for the development of vaccines and antiviral agents. J Virol 79, 3289–3296. DOI:10.1128/JVI.79.6.3289-3296.2005

Khoury, D.S., Cromer, D., Reynaldi, A., Schlub, T.E., Wheatley, A.K., Juno, J.A., Subbarao, K., Kent, S.J., Triccas, J.A., and Davenport, M.P. (2021). Neutralizing antibody levels are highly predictive of immune protection from symptomatic SARS-CoV-2 infection. Nat Med. DOI:10.1038/s41591-021-01377-8

Khoury, D.S., Wheatley, A.K., Ramuta, M.D., Reynaldi, A., Cromer, D., Subbarao, K., O’Connor, D.H., Kent, S.J., and Davenport, M.P. (2020). Measuring immunity to SARS-CoV-2 infection: comparing assays and animal models. Nat Rev Immunol 20, 727–738. DOI:10.1038/s41577-020-00471-1

Korber, B., Fischer, W.M., Gnanakaran, S., Yoon, H., Theiler, J., Abfalterer, W., Hengartner, N., Giorgi, E.E., Bhattacharya, T., Foley, B., et al. (2020). Tracking Changes in SARS-CoV-2 Spike: Evidence that D614G Increases Infectivity of the COVID-19 Virus. Cell 182, 812–827 e819. DOI:10.1016/j.cell.2020.06.043

Krammer, F. (2021). A correlate of protection for SARS-CoV-2 vaccines is urgently needed. Nat Med 27, 1147–1148. DOI:10.1038/s41591-021-01432-4

Kristiansen, P.A., Page, M., Bernasconi, V., Mattiuzzo, G., Dull, P., Makar, K., Plotkin, S., and Knezevic, I. (2021). WHO International Standard for anti-SARS-CoV-2 immunoglobulin. Lancet 397, 1347–1348. DOI:10.1016/S0140-6736(21)00527-4

Levine-Tiefenbrun, M., Yelin, I., Alapi, H., Katz, R., Herzel, E., Kuint, J., Chodick, G., Gazit, S., Patalon, T., and Kishony, R. (2021). Viral loads of Delta-variant SARS-CoV-2 breakthrough infections after vaccination and booster with BNT162b2. Nat Med. DOI:10.1038/s41591-021-01575-4

Liu, C., Ginn, H.M., Dejnirattisai, W., Supasa, P., Wang, B., Tuekprakhon, A., Nutalai, R., Zhou, D., Mentzer, A.J., Zhao, Y., et al. (2021a). Reduced neutralization of SARS-CoV-2 B.1.617 by vaccine and convalescent serum. Cell 184, 4220–4236 e4213. DOI:10.1016/j.cell.2021.06.020

Liu, C., Mendonca, L., Yang, Y., Gao, Y., Shen, C., Liu, J., Ni, T., Ju, B., Liu, C., Tang, X., et al. (2020). The Architecture of Inactivated SARS-CoV-2 with Postfusion Spikes Revealed by Cryo-EM and Cryo-ET. Structure 28, 1218–1224 e1214. DOI:10.1016/j.str.2020.10.001

Liu, Y., and Rocklov, J. (2021). The reproductive number of the Delta variant of SARS-CoV-2 is far higher compared to the ancestral SARS-CoV-2 virus. J Travel Med 28. DOI:10.1093/jtm/taab124

Liu, Y., Soh, W.T., Kishikawa, J.I., Hirose, M., Nakayama, E.E., Li, S., Sasai, M., Suzuki, T., Tada, A., Arakawa, A., et al. (2021b). An infectivity-enhancing site on the SARS-CoV-2 spike protein targeted by antibodies. Cell 184, 3452–3466 e3418. DOI:10.1016/j.cell.2021.05.032

Lopez Bernal, J., Andrews, N., Gower, C., Gallagher, E., Simmons, R., Thelwall, S., Stowe, J., Tessier, E., Groves, N., Dabrera, G., et al. (2021). Effectiveness of Covid-19 Vaccines against the B.1.617.2 (Delta) Variant. N Engl J Med 385, 585–594. DOI:10.1056/NEJMoa2108891

Luo, C.H., Morris, C.P., Sachithanandham, J., Amadi, A., Gaston, D., Li, M., Swanson, N.J., Schwartz, M., Klein, E.Y., Pekosz, A., et al. (2021). Infection with the SARS-CoV-2 Delta Variant is Associated with Higher Infectious Virus Loads Compared to the Alpha Variant in both Unvaccinated and Vaccinated Individuals. medRxiv. DOI:10.1101/2021.08.15.21262077

Millet, J.K., and Whittaker, G.R. (2015). Host cell proteases: Critical determinants of coronavirus tropism and pathogenesis. Virus Res 202, 120–134. DOI:10.1016/j.virusres.2014.11.021

Mittal, A., Manjunath, K., Ranjan, R.K., Kaushik, S., Kumar, S., and Verma, V. (2020). COVID-19 pandemic: Insights into structure, function, and hACE2 receptor recognition by SARS-CoV-2. PLoS Pathog 16, e1008762. DOI:10.1371/journal.ppat.1008762

Miyakawa, K., Jeremiah, S.S., Ohtake, N., Matsunaga, S., Yamaoka, Y., Nishi, M., Morita, T., Saji, R., Nishii, M., Kimura, H., et al. (2020). Rapid quantitative screening assay for SARS-CoV-2 neutralizing antibodies using HiBiT-tagged virus-like particles. J Mol Cell Biol 12, 987–990. DOI:10.1093/jmcb/mjaa047

Niessl, J., Sekine, T., and Buggert, M. (2021). T cell immunity to SARS-CoV-2. Semin Immunol, 101505. DOI:10.1016/j.smim.2021.101505

Niu, L., Wittrock, K.N., Clabaugh, G.C., Srivastava, V., and Cho, M.W. (2021). A Structural Landscape of Neutralizing Antibodies Against SARS-CoV-2 Receptor Binding Domain. Front Immunol 12, 647934. DOI:10.3389/fimmu.2021.647934

Ou, X., Liu, Y., Lei, X., Li, P., Mi, D., Ren, L., Guo, L., Guo, R., Chen, T., Hu, J., et al. (2020). Characterization of spike glycoprotein of SARS-CoV-2 on virus entry and its immune cross-reactivity with SARS-CoV. Nat Commun 11, 1620. DOI:10.1038/s41467-020-15562-9

Papa, G., Mallery, D.L., Albecka, A., Welch, L.G., Cattin-Ortola, J., Luptak, J., Paul, D., McMahon, H.T., Goodfellow, I.G., Carter, A., et al. (2021). Furin cleavage of SARS-CoV-2 Spike promotes but is not essential for infection and cell-cell fusion. PLoS Pathog 17, e1009246. DOI:10.1371/journal.ppat.1009246

Piccoli, L., Park, Y.J., Tortorici, M.A., Czudnochowski, N., Walls, A.C., Beltramello, M., Silacci-Fregni, C., Pinto, D., Rosen, L.E., Bowen, J.E., et al. (2020). Mapping Neutralizing and Immunodominant Sites on the SARS-CoV-2 Spike Receptor-Binding Domain by Structure-Guided High-Resolution Serology. Cell 183, 1024–1042 e1021. DOI:10.1016/j.cell.2020.09.037

Planas, D., Veyer, D., Baidaliuk, A., Staropoli, I., Guivel-Benhassine, F., Rajah, M.M., Planchais, C., Porrot, F., Robillard, N., Puech, J., et al. (2021). Reduced sensitivity of SARS-CoV-2 variant Delta to antibody neutralization. Nature 596, 276–280. DOI:10.1038/s41586-021-03777-9

Romero-Olmedo, A.J., Schulz, A.R., Hochstätter, S., Gupta, D.D., Virta, I., Hirseland, H., Staudenraus, D., Camara, B., Münch, C., Hefter, V., et al. (2021). Induction of robust cellular and humoral immunity against SARS-CoV-2 after a third dose of BNT162b2 vaccine in previously unresponsive elderly. Nat Microbiology in press.

Shang, J., Wan, Y., Luo, C., Ye, G., Geng, Q., Auerbach, A., and Li, F. (2020). Cell entry mechanisms of SARS-CoV-2. Proc Natl Acad Sci U S A 117, 11727–11734. DOI:10.1073/pnas.2003138117

Shanmugaraj, B., Siriwattananon, K., Wangkanont, K., and Phoolcharoen, W. (2020). Perspectives on monoclonal antibody therapy as potential therapeutic intervention for Coronavirus disease-19 (COVID-19). Asian Pac J Allergy Immunol 38, 10–18. DOI:10.12932/AP-200220-0773

Sheikh, A., McMenamin, J., Taylor, B., Robertson, C., Public Health, S., and the, E.I.I.C. (2021). SARS-CoV-2 Delta VOC in Scotland: demographics, risk of hospital admission, and vaccine effectiveness. Lancet 397, 2461–2462. DOI:10.1016/S0140-6736(21)01358-1

Singanayagam, A., Hakki, S., Dunning, J., Madon, K.J., Crone, M.A., Koycheva, A., Derqui-Fernandez, N., Barnett, J.L., Whitfield, M.G., Varro, R., et al. (2021). Community transmission and viral load kinetics of the SARS-CoV-2 delta (B.1.617.2) variant in vaccinated and unvaccinated individuals in the UK: a prospective, longitudinal, cohort study. Lancet Infect Dis. DOI:10.1016/S1473-3099(21)00648-4

Siu, Y.L., Teoh, K.T., Lo, J., Chan, C.M., Kien, F., Escriou, N., Tsao, S.W., Nicholls, J.M., Altmeyer, R., Peiris, J.S., et al. (2008). The M, E, and N structural proteins of the severe acute respiratory syndrome coronavirus are required for efficient assembly, trafficking, and release of virus-like particles. J Virol 82, 11318–11330. DOI:10.1128/JVI.01052-08

Somiya, M., and Kuroda, S. (2021). Real-Time Luminescence Assay for Cytoplasmic Cargo Delivery of Extracellular Vesicles. Anal Chem 93, 5612–5620. DOI:10.1021/acs.analchem.1c00339

Stevens, C.S., Oguntuyo, K.Y., and Lee, B. (2021). Proteases and variants: context matters for SARS-CoV-2 entry assays. Curr Opin Virol 50, 49–58. DOI:10.1016/j.coviro.2021.07.004

Syed, A.M., Taha, T.Y., Tabata, T., Chen, I.P., Ciling, A., Khalid, M.M., Sreekumar, B., Chen, P.Y., Hayashi, J.M., Soczek, K.M., et al. (2021). Rapid assessment of SARS-CoV-2 evolved variants using virus-like particles. Science, eabl6184. DOI:10.1126/science.abl6184

Tortorici, M.A., Beltramello, M., Lempp, F.A., Pinto, D., Dang, H.V., Rosen, L.E., McCallum, M., Bowen, J., Minola, A., Jaconi, S., et al. (2020). Ultrapotent human antibodies protect against SARS-CoV-2 challenge via multiple mechanisms. Science 370, 950–957. DOI:10.1126/science.abe3354

Turonova, B., Sikora, M., Schurmann, C., Hagen, W.J.H., Welsch, S., Blanc, F.E.C., von Bulow, S., Gecht, M., Bagola, K., Horner, C., et al. (2020). In situ structural analysis of SARS-CoV-2 spike reveals flexibility mediated by three hinges. Science 370, 203–208. DOI:10.1126/science.abd5223

Ujike, M., Huang, C., Shirato, K., Makino, S., and Taguchi, F. (2016). The contribution of the cytoplasmic retrieval signal of severe acute respiratory syndrome coronavirus to intracellular accumulation of S proteins and incorporation of S protein into virus-like particles. J Gen Virol 97, 1853–1864. DOI:10.1099/jgv.0.000494

von Rhein, C., Scholz, T., Henss, L., Kronstein-Wiedemann, R., Schwarz, T., Rodionov, R.N., Corman, V.M., Tonn, T., and Schnierle, B.S. (2021). Comparison of potency assays to assess SARS-CoV-2 neutralizing antibody capacity in COVID-19 convalescent plasma. J Virol Methods 288, 114031. DOI:10.1016/j.jviromet.2020.114031

World Health Organization (2020). Clinical management of COVID-19: interim guidance, 27 May 2020 (Geneva: World Health Organization).

Wu, F., Zhao, S., Yu, B., Chen, Y.M., Wang, W., Song, Z.G., Hu, Y., Tao, Z.W., Tian, J.H., Pei, Y.Y., et al. (2020). A new coronavirus associated with human respiratory disease in China. Nature 579, 265–269. DOI:10.1038/s41586-020-2008-3

Yamamoto, M., Du, Q., Song, J., Wang, H., Watanabe, A., Tanaka, Y., Kawaguchi, Y., Inoue, J.I., and Matsuda, Z. (2019). Cell-cell and virus-cell fusion assay-based analyses of alanine insertion mutants in the distal alpha9 portion of the JRFL gp41 subunit from HIV-1. J Biol Chem 294, 5677–5687. DOI:10.1074/jbc.RA118.004579

Yang, R., Huang, B., A, R., Li, W., Wang, W., Deng, Y., and Tan, W. (2020). Development and effectiveness of pseudotyped SARS-CoV-2 system as determined by neutralizing efficiency and entry inhibition test in vitro. Biosaf Health 2, 226–231. DOI:10.1016/j.bsheal.2020.08.004

Yao, H., Song, Y., Chen, Y., Wu, N., Xu, J., Sun, C., Zhang, J., Weng, T., Zhang, Z., Wu, Z., et al. (2020). Molecular Architecture of the SARS-CoV-2 Virus. Cell 183, 730–738 e713. DOI:10.1016/j.cell.2020.09.018

Zahradnik, J., Marciano, S., Shemesh, M., Zoler, E., Harari, D., Chiaravalli, J., Meyer, B., Rudich, Y., Li, C., Marton, I., et al. (2021). SARS-CoV-2 variant prediction and antiviral drug design are enabled by RBD in vitro evolution. Nat Microbiol 6, 1188–1198. DOI:10.1038/s41564-021-00954-4

Zhang, L., Jackson, C.B., Mou, H., Ojha, A., Peng, H., Quinlan, B.D., Rangarajan, E.S., Pan, A., Vanderheiden, A., Suthar, M.S., et al. (2020). SARS-CoV-2 spike-protein D614G mutation increases virion spike density and infectivity. Nat Commun 11, 6013. DOI:10.1038/s41467-020-19808-4

Zohar, T., and Alter, G. (2020). Dissecting antibody-mediated protection against SARS-CoV-2. Nat Rev Immunol 20, 392–394. DOI:10.1038/s41577-020-0359-5

