## Supplementary Table S1 for "Quantitation of SARS-CoV-2 neutralizing antibodies with a virus-free, authentic test"

**Supplementary Table S1. Individual data of analyzed human serum samples.**

| | | | | | | | | | | | VLPNT | | | cVNT | |
|---|---|---|---|---|---|---|---|---|---|---|---|---|---|---|---|
| | | | | | | | Vaccination scheme | | | | D614G | | B.1.617.2 | D614G | |
| Group | ID | Sex | Age group | Sampling | COVID-19 severity | RT-qPCR | Prime | Boost | d p. PCR[a] | d p. Boost[b] | $VLPN_{50}$ [1: ] | $Titer_{50}$[c] [IU/mL] | $VLPN_{50}$ [1: ] | $VNT_{100}$[d] [1: ] | $Titer_{100}$ [IU/mL] |
| COVID-19 patients | A1 | f | 31-35 | Jun 2020 | asymptomatic | + | | | 104 | | 49 | 76 | | 8 | 21 |
| | A2 | f | 61-65 | Jun 2020 | moderate dis. | + | | | 54 | | 352 | 546 | | 81 | 215 |
| | A3 | m | 56-60 | Jun 2020 | asymptomatic | + | | | 79 | | 18 | 28 | | 8 | 21 |
| | A4 | m | 31-35 | Jun 2020 | severe dis. | + | | | 76 | | >2000 | >3101 | | >1024 | >2716 |
| | A5 | m | 56-60 | May 2020 | mild dis. | + | | | 17 | | 26 | 40 | | †4 | †11 |
| | A6 | m | 36-40 | May 2020 | mild dis. | + | | | 70 | | 61 | 95 | | 16 | 42 |
| | A7 | m | 71-75 | May 2020 | severe dis. | + | | | 4 | | 89 | 138 | | †4 | †11 |
| | A8 | m | 51-55 | May 2020 | moderate dis. | + | | | 57 | | >2000 | >3101 | | >1024 | >2716 |
| | A9 | m | 81-85 | May 2020 | severe dis. | + | | | 53 | | 89 | 138 | | 25 | 66 |
| | A10 | m | 56-60 | May 2020 | mild dis. | + | | | 11 | | 58 | 90 | | †4 | †11 |
| | A11 | m | 56-60 | May 2020 | moderate dis. | + | | | 38 | | 340 | 527 | | 102 | 271 |
| | A12 | m | 71-75 | May 2020 | critical dis. | + | | | 13 | | 329 | 510 | | 203 | 538 |
| | B1 | m | 61-65 | May 2020 | asymptomatic | + | | | 15 | | 142 | 220 | | 81 | 215 |
| | B2 | m | 51-55 | May 2020 | moderate dis. | + | | | 26 | | 645 | 1000 | | >1024 | >2716 |
| | B3 | m | 86-90 | Apr 2020 | moderate dis. | + | | | 9 | | 426 | 660 | | 323 | 857 |
| | B4 | f | 31-35 | May 2020 | asymptomatic | + | | | 54 | | 61 | 95 | | †4 | †11 |
| | B5 | m | 81-85 | Apr 2020 | severe dis. | + | | | 22 | | 46 | 71 | | 40 | 106 |
| | B6 | m | 81-85 | Apr 2020 | severe dis. | + | | | 18 | | 29 | 45 | | 10 | 27 |
| | B7 | m | 56-60 | Apr 2020 | severe dis. | + | | | 12 | | 1527 | 2367 | | 512 | 1358 |
| | B8 | m | 86-90 | Apr 2020 | severe dis. | + | | | 2 | | 48 | 74 | | †4 | †11 |
| | B9 | m | 86-90 | Apr 2020 | critical dis. | + | | | 2 | | 44 | 68 | | †4 | †11 |
| | B10 | m | 56-60 | Apr 2020 | severe dis. | + | | | 8 | | 1250 | 1938 | | 645 | 1711 |
| | B11 | m | 26-30 | Apr 2020 | moderate dis. | + | | | 1 | | 125 | 194 | | 81 | 215 |
| COVID-19 vaccinees | S001 | f | 91-95 | Feb 2021 | | | BNT162b2 | BNT162b2 | | 24 | 100 | 155 | 37 | | |
| | S005 | m | 26-30 | May 2021 | | | AZD1222 | BNT162b2 | | 13 | 82 | 113 | 47 | | |
| | S008 | m | 56-60 | May 2021 | | | AZD1222 | BNT162b2 | | 14 | 27 | 73 | 24 | | |
| | S011 | f | 26-30 | Jul 2021 | | | mRNA-1273 | mRNA-1273 | | 19 | 352 | 860 | 178 | | |
| | S012 | m | 66-70 | Jul 2021 | | | AZD1222 | mRNA-1273 | | 15 | 99 | 242 | 81 | | |
| | S014 | m | 66-70 | Jul 2021 | | | AZD1222 | AZD1222 | | 37 | 63 | 154 | 45 | | |
| | S015 | f | 31-35 | Jul 2021 | | | mRNA-1273 | mRNA-1273 | | 13 | 267 | 654 | 254 | | |
| | S016 | f | 56-60 | Jul 2021 | | | BNT162b2 | BNT162b2 | | 56 | 38 | 91 | 30 | | |
| | S025 | f | 66-70 | Aug 2021 | | | AZD1222 | mRNA-1273 | | 22 | 254 | 697 | 104 | | |
| | S026 | m | 66-70 | Aug 2021 | | | AZD1222 | mRNA-1273 | | 15 | 92 | 162 | 81 | | |
| | S027 | f | 56-60 | Aug 2021 | | | BNT162b2 | BNT162b2 | | 14 | 267 | 734 | 105 | | |
| | S033 | f | 56-60 | Sep 2021 | | | BNT162b2 | BNT162b2 | | 29 | 69 | 162 | 36 | | |
| | S034 | m | 56-60 | Sep 2021 | | | BNT162b2 | BNT162b2 | | 90 | 40 | 94 | 17 | | |
| Healthy, naive donors | S003 | m | 21-25 | Aug 2019 | | | | | | | 5 | 8 | | | |
| | S006 | m | 56-60 | Oct 2017 | | | | | | | 14 | 22 | | | |
| | S009 | f | 26-30 | May 2019 | | | | | | | 0 | 0 | | | |
| | S018 | f | 26-30 | Oct 2017 | | | | | | | 16 | 25 | | | |
| | S019 | f | 66-70 | Nov 2017 | | | | | | | 0 | 0 | | | |
| | S020 | f | 31-35 | Nov 2017 | | | | | | | 0 | 0 | | | |
| | S021 | f | 41-45 | Dec 2017 | | | | | | | 0 | 0 | | | |
| | S022 | f | 51-55 | Dec 2017 | | | | | | | 0 | 0 | | | |
| | S023 | f | 31-35 | Oct 2017 | | | | | | | 0 | 0 | | | |
| | S024 | m | na | Dec 2017 | | | | | | | 6 | 9 | | | |
| | S028 | m | 26-30 | Mar 2019 | | | | | | | 0 | 0 | | | |
| | S029 | f | 31-35 | Mar 2019 | | | | | | | 10 | 16 | | | |
| NIBSC 20/136 | WHO | na | na | 2020 | | | | | | | 645 | *1000 | | 377 | *1000 |
| | WHO | na | na | 2020 | | | | | | | 409 | *1000 | | | |
| | WHO | na | na | 2020 | | | | | | | 417 | *1000 | | | |
| | WHO | na | na | 2020 | | | | | | | 364 | *1000 | | | |
| | WHO | na | na | 2020 | | | | | | | 567 | *1000 | | | |
| | WHO | na | na | 2020 | | | | | | | 426 | *1000 | | | |

m, male; f, female; na, not available; dis., disease; d, days; p., post; *, per WHO definition; †, below limit of detection

[a]Days between first positive SARS-CoV-2 RT-qPCR report and sampling

[b]Days between booster dose and sampling

[c]$VLPN_{50}$ titer normalized to the WHO reference serum's within-run performance

[d]Reciprocal geometric mean titer (GMT) from the highest serum dilution displaying 100% reduction of CPE based on three replicates
